# Systems Immunology of Long Covid: Insights from the STOP-PASC Clinical Trial

**DOI:** 10.64898/2025.12.04.25341650

**Authors:** Evan Maestri, Woo Joo Kwon, Hong Zheng, Tyler Prestwood, Haley Hedlin, Jane W. Liang, Holly McCann, Blake Shaw, Lu Tian, Ben Jones, Rufei Lu, Graham Wiley, Emily Haraguchi, Oliver Wirz, Jumana Afaghani, Brandon Lam, Dlovan F. D Mahmood, Nicole A. Phillips, Martha M.S. Sim, Jeremy P. Wood, James R. Heath, Scott D. Boyd, Joel Guthridge, Upinder Singh, Hector Bonilla, Prasanna Jagannathan, PJ Utz, Linda N. Geng, Purvesh Khatri

## Abstract

**Background:** Post Acute Sequelae of COVID-19 (PASC), also referred to as Long COVID, is an infection-associated chronic syndrome with heterogenous symptom profiles that occurs in a subset of people following SARS-CoV-2 infection. Despite proposed viral persistence mechanisms, no therapeutic benefit was observed in two randomized placebo-controlled trials of nirmatrelvir/ritonavir (NMV/r) in adults with Long COVID, including the Selective Trial of Paxlovid for PASC (STOP-PASC) and PAX LC. This systems immunology analysis aimed to characterize immune profiles of participants during clinical trial intervention, identify biomarkers associated with patient-reported outcomes, and investigate potential mechanisms underlying Long COVID.

**Methods:** We performed comprehensive immunological profiling of 152 STOP-PASC trial participants using plasma proteomics (Olink® Explore HT 5400 panel), autoantigen arrays, viral serology, and microclot assays at baseline, day 15, and week 10. We assessed associations between immune features and patient-reported outcomes. We also conducted meta-analysis of nine independent Long COVID proteomics cohorts (n=590 total samples) to identify conserved inflammatory signatures.

**Results:** NMV/r treatment at day 15 compared with baseline induced transient changes in plasma proteins that normalized by week 10, primarily impacting myeloid cell/monocyte, lysosome, and complement activation pathways. Cardiovascular symptoms were negatively associated with SARS-CoV-2 antibody levels at baseline. No widespread differences in autoantibody profiles, Epstein-Barr virus (EBV) reactivation, or microclotting were observed between STOP-PASC Long COVID participants, pre-pandemic controls, and individuals without Long COVID. Meta-analysis of publicly available Olink® data from Long COVID cohorts identified a conserved 60-protein Long COVID Signature (LCS) score revealing multi-compartment immune activation involving monocyte, neutrophil, and T/NK cell modules.

**Conclusions:** These findings advance our understanding of Long COVID immunology and may help direct future proteomic biomarker endpoints for Long COVID clinical trials.

## Introduction

Long COVID (LC) is an infection-associated chronic condition estimated to impact at least 8.4% of adults in the United States population^1–3^. Its clinical presentation and disease trajectory is extremely heterogeneous, with common symptoms including post-exertional malaise, fatigue, brain fog, dizziness, and shortness of breath^4^. The multi-system, relapsing and remitting nature of LC, and its complex debilitating symptomatology that impacts multiple organ systems can impose significant burden on quality of life^5^. Several molecular mechanisms have been postulated to explain LC, including persistent inflammation and/or autoimmunity, reactivation of viruses including Epstein-Barr Virus (EBV) or other herpesviruses, and persistence of SARS-CoV-2 in the gut, brain, or other tissues^6^. Since each etiology would be treated through different approaches, it is critical to identify actionable biomarkers to enable more targeted therapies.

Immune profiling studies in LC have examined cellular, cytokine, and antibody responses to better understand its underlying pathophysiology^7–9^. Klein et al. systematically analyzed potential LC biomarkers identifying differences in circulating immune cell populations, elevated antibody responses to herpesviruses antigens (EBV and varicella-zoster virus), and little differences in autoantibodies to the human exoproteome^7^. Yin and colleagues identified T cell dysregulation in LC, including increased expression of exhaustion markers in SARS-CoV-2-specific CD8+ T cells^8^. Su et al. used multi-omics longitudinal investigation to show that early immunological parameters at initial COVID-19 diagnosis, such as pre-existing type 2 diabetes, SARS-CoV-2 RNAemia, EBV viremia, and certain autoantibodies could help anticipate which patients may develop persistent symptoms^9^. In addition, proteomics studies have revealed increased complement activation and thromboinflammation, ongoing neutrophil activity, and persistent inflammation^10–12^. Despite these advances, our current understanding of LC mechanisms, immunological diagnostics, and effective therapeutic approaches remains incomplete. To aid millions currently struggling with post-acute viral syndromes, and for enhanced future pandemic preparedness, we need to better understand the complexity of the immune system in LC pathogenesis to reduce disability burden.

Persistence of SARS-COV-2 following initial infection was postulated by several groups and may be a cause of LC, motivating randomized clinical trials of the oral antiviral nirmatrelvir-ritonavir (NMV/r) in adults with LC. We recently conducted a randomized controlled trial of the Selective Trial of Paxlovid for PASC (STOP-PASC, NCT05576662), where individuals with LC were randomized to receive NMV/r vs. placebo^13^. Although there were no differences in patient-reported outcomes comparing NMV/r to placebo/r (PBO/r)^13^, given the high individual heterogeneity among study participants it was unclear if immunological profiles varied during therapy.

This exploratory analysis aimed to evaluate the following objectives. First, we investigated potential peripheral blood biological biomarkers of LC in STOP-PASC participants treated with NMV/r versus PBO/r, including identifying treatment-specific changes in the plasma proteome during therapy. Second, we sought to determine whether patient-reported outcome measures were associated with immune profiles. Third, we characterize anti- viral IgG antibody responses and autoantibody profiles in LC. Lastly, we investigated conserved proteomics signatures distinguishing LC patients using multi-cohort meta-analysis of public data. Overall, we present the immunological findings of the STOP-PASC trial, which yields insights into potential biomarkers and LC mechanisms, and offers valuable context for the future development of therapeutic interventions.

## Results

### Study Population

The STOP-PASC clinical trial and its digital biometric substudy have been described previously^13–15^. Briefly, 155 participants with Long COVID (LC) were randomized 2:1 to receive either nirmatrelvir-ritonavir or placebo- ritonavir for 15 days and n=152 participants (n=100 NMV/r, n=52 PBO/r) with sufficient post-baseline biological samples were included for systems immunology profiling analysis (**Figure 1**). Trial demographic participant characteristics were previously described^13^. For comparative analyses against the STOP-PASC LC participants, two control cohorts were also included in the immune profiling. The healthy pre-pandemic controls had a mean (SD) age of 43.14 (7.94), with 22 (52.4%) female participants (**Table S1**). The LC negative (LC^neg^) population (individuals without LC) had a mean (SD) age of 48.12 (13.08), with 22 (52.4%) female participants. The standardized mean differences (SMD) indicated the differences between STOP PASC participants and control groups were small for age (SMD=0.32), sex (SMD=0.1), ethnicity (SMD=0.29), and race (SMD=0.4).

**Figure 1.**
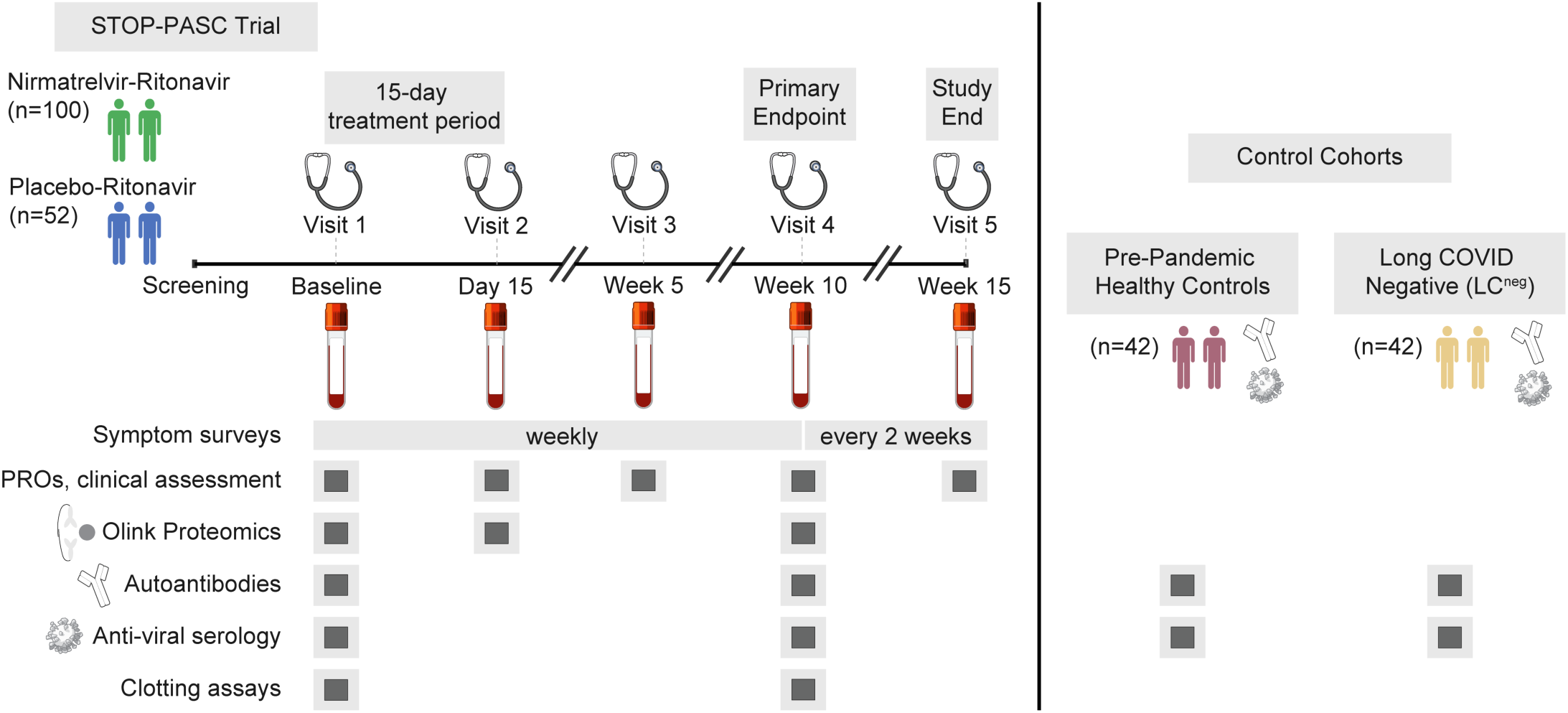
Study design and immunological profiling framework. Analysis of immune responses in participants (n=152) in the STOP-PASC clinical trial of NMV/r treatment vs. PBO/r in Long COVID (LC). The study duration was 15 weeks with 5 in-person visits conducted at baseline, day 15, and at weeks 5, 10, and 15. At each time point, symptom assessment, orthostatic blood pressure measurements, vitals, stool collection, and nasopharyngeal swabs for SARS-CoV-2 testing were performed. Blood collected at days 0, 15, and week 10 were evaluated by immunologic assays including plasma proteomics (Olink®), custom microbead-based autoantigen arrays, SARS-CoV-2-specific antibody measurements, and clotting assays. PRO: patient-reported outcome. Control cohorts were used for comparative analysis of autoantibodies and anti-viral serology.

### Magnitude of anti-viral IgG in Long COVID (LC)

Anti-spike IgG responses and their trajectories have been postulated to differ in patients with LC compared with individuals with acute infections who fully recover^16^. Moreover, prior infections with related seasonal coronaviruses such as OC43 may alter the nature and duration of the immune response to SARS-CoV-2 infection^17^. Using two different methodologies we measured IgG directed against multiple different viruses including SARS-CoV-2 variants, other pathogenic coronaviruses including MERS, SARS-CoV, and seasonal coronaviruses in STOP-PASC participants. We quantified SARS-CoV-2-specific immunoglobulin G (IgG) responses with multiplexed electrochemiluminescence (ECL) assays (Meso Scale Discovery, MSD). We detected antibodies against multiple SARS-CoV-2 proteins including the Spike (S) protein, the Receptor Binding Domain (RBD), and N-terminal domain (NTD), across several variants including Omicron and Delta, as well as other unrelated respiratory viruses such as influenza and respiratory syncytial virus (RSV). We also measured IgG antibodies against additional pathogens including Epstein-Barr Virus (EBV), cytomegalovirus (CMV), and seasonal coronaviruses (229E alpha coronavirus, HKU1 beta coronavirus, OC43 beta coronavirus) using a custom microbead protein array (**Table S2**). As expected, healthy uninfected pre-pandemic control samples lacked antibodies against Spike protein (**Figure S1**). Anti-S, anti-RBD, and anti-NTD were significantly different between STOP-PASC LC participants at baseline (LC^BL^) and LC^neg^ (individuals without LC) (**Figure 2A, S2**). Anti-N IgG levels, specific for N protein not encoded in the vaccine, did not differ between groups. By Week 10 of the trial, NTD-specific IgG, RBD-specific and Spike-specific IgG decreased >1.4-fold from baseline levels (**Figure 2B,C and S3**). IgG antibodies against Herpesvirus proteins including EBV Viral Capsid Antigen (VCA), EBV Nuclear Antigen 1 (EBNA-1), Early Antigen (EA) and CMV glycoprotein B (gB) did not differ between STOP-PASC LC^BL^ participants and LC^neg^ individuals (**Figure 2D, S4**). IgG titers against the seasonal human coronaviruses 229E, HKU1, and OC43 were not significantly different between STOP-PASC LC^BL^ participants and LC^neg^ after multiple hypothesis correction; however, reactivities were greatly increased compared to pre-pandemic controls (**Figure 2D**). Next, we assessed if any viral serology measures from either MSD or microbead platforms at baseline in STOP-PASC LC participants were associated with the severity of the core patient-reported outcomes (PROs) reported by the participants in the STOP-PASC study. We assessed associations using proportional odds logistic regression models fit to the Likert scale ordinal severity level of each core symptom at baseline. The outcomes assessed were the six core symptoms (fatigue, brain fog, dyspnea, body aches, heart, stomach) and an additional post-exertional malaise symptom. Of the PROs tested, negative associations between heart symptom severity and antibody levels were observed (**Figure S5**). Individuals with no or fewer heart symptoms had higher antibody levels against a subset of SARS-CoV-2 proteins, such as Delta variant AY.4, CoV- 2 Spike, CoV-2 NTD, and CoV-2 RBD (**Figure 2E,F**).

**Figure 2.**
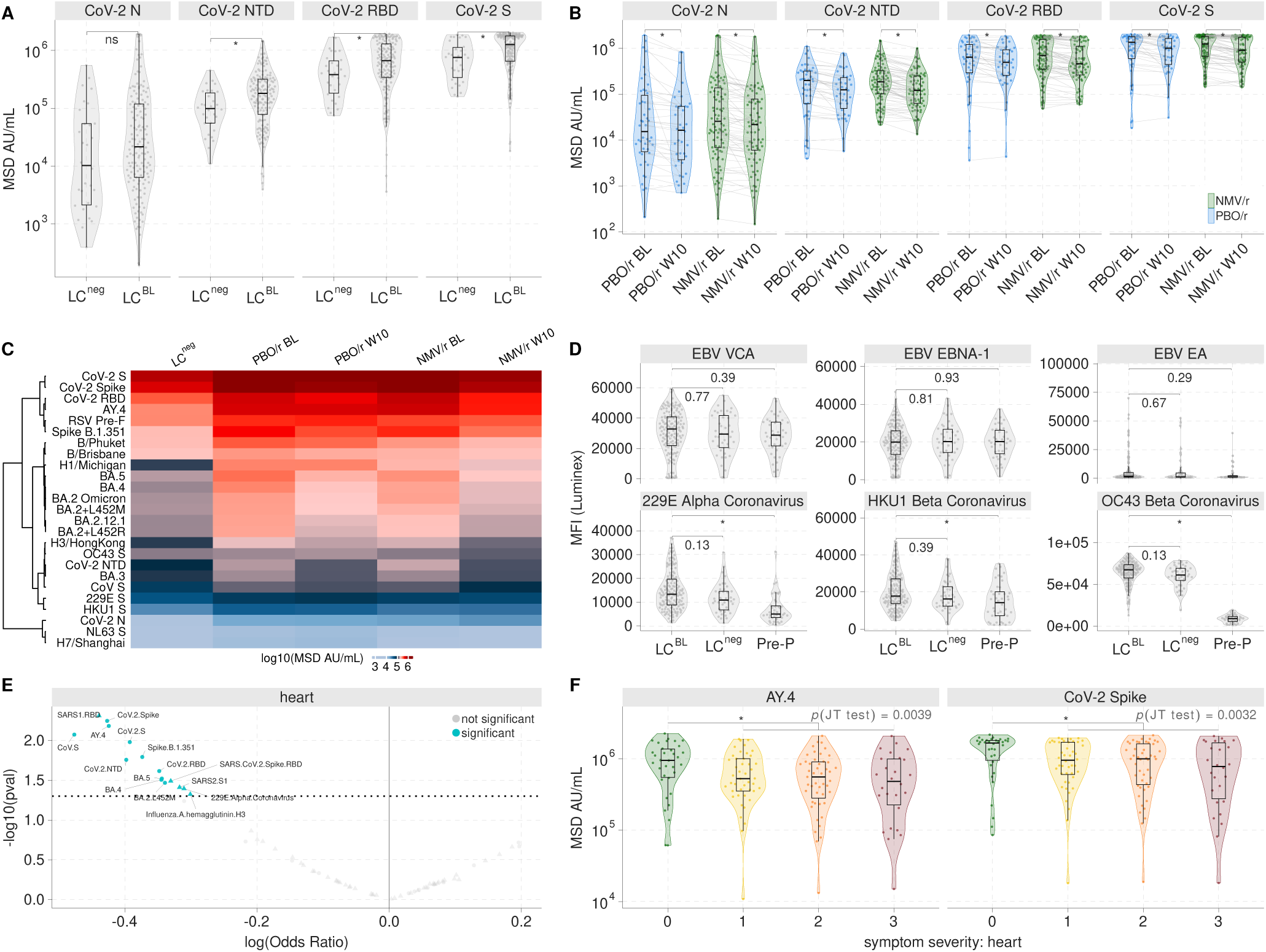
Viral serological responses and associations with LC patient-reported outcomes. (A) Anti–SARS-CoV-2 N, N-terminal domain (NTD), Spike (S), and receptor-binding domain (RBD) IgG antibody levels expressed in MSD arbitrary units (AU/mL). Statistical test: Wilcoxon rank-sum test with FDR correction. STOP-PASC LC participants at baseline (LC^BL^); Individuals without LC (LC^neg^). (B) Longitudinal antibody dynamics between BL and week 10 in MSD AU/mL (blue: PBO/r, green: NMV/r). Statistical test: pairwise Wilcoxon rank-sum test with Bonferroni correction. (C) Heatmap of anti–SARS-CoV-2, seasonal coronaviruses, and influenza IgG antibody levels in LC^neg^, PBO/r and NMV/r at baseline and week 10. The color scale represents the median values of log10 MSD AU/mL concentrations. (D) Herpesvirus and seasonal coronavirus IgG antibody levels in LC^neg^, LC^BL^, and pre-pandemic (Pre-P) samples measured using a custom, multiplexed, microbead protein array. Statistical test: Wilcoxon rank-sum test with FDR correction (Benjamini-Hochberg). (E) Association between antibody level and PRO severity in STOP-PASC at baseline as estimated by proportional odds logistic regression. The color indicates significance with *P*<0.05 (not corrected for multiple testing). Circles are measurements from MSD and triangles are from microbead assays. (F) Antibody levels for AY.4 and CoV-2 Spike and their association with heart symptom severity on a Likert scale (where 0 is none, 1 mild, 2 moderate, 3 severe). Jonckheere-Terpstra (JT) trend test p-value indicates significance of the trend over severity.

### Longitudinal autoantibody and clotting profiling of Long COVID

Prior reports have identified elevated autoantibodies (AAbs) and increased risk for new-onset autoimmune disease following acute SARS-CoV-2 infection^18,19^. We characterized AAb profiles in STOP-PASC participants with LC at baseline and week 10 (100 NMV/r, 52 PBO/r), 42 healthy uninfected pre-pandemic controls, and 42 LC^neg^ using a custom, 76-plex protein array (**Table S3**) consisting of connective tissue disease antigens, cytokines, chemokines, and growth factors to measure anti-cytokine antibodies (ACA). There were no differences in plasma AAb profiles between any of the groups (**Figure S6A**). The prevalence of AAb was similar in the NMV/r and PBO/r arms. A subset of LC participants had reactivities at baseline against CENP A (7.8%, 12/153), fibrillarin (4.5%, 7/153), and PDC-E2 (4.5%, 7/153) which remained at week 10 (**Figure S6B-E**), the clinical significance of which is unclear.

Acute COVID, as well as LC, is associated with abnormalities in thrombosis, including development of pathogenic anti-phospholipid antibodies, venous thrombosis, pulmonary embolism, endothelial activation, and microclots, all of which can be caused by dysregulation of components of the clotting cascade^20^. Microclots were detected only in a small subset of STOP-PASC LC participants at baseline and the standardized mean differences (SMD) suggest that differences between groups are generally small (**Figure S7, Table S4**).

### Temporal changes in the plasma proteome during NMV/r treatment

To gain insights into the effect of NMV/r on the plasma proteome, we measured the concentration of 5400 analytes using Olink® in 421 total samples from STOP-PASC LC participants at baseline (100 NMV/r, 52 PBO/r), day 15 (94 NMV/r, 48 PBO/r), and week 10 (85 NMV/r, 42 PBO/r) timepoints. In the NMV/r group, there were 94 statistically significant proteins (adjusted *P*≤0.05) on day 15 compared to baseline, whereas only 9 proteins were significantly different in the PBO/r group compared to baseline (**Figure 3A,B, Table S5**), demonstrating impact of NMV/r treatment. At day 15, CD14, Cathepsin Z (CTSZ), Hyaluronic acid binding protein 2 (HABP2), and Complement C1q tumor necrosis factor-related protein 1 (C1QTNF1) were differentially upregulated specific to the NMV/r group (**Figure 3C**). In addition, multiple coagulation factors including X (F10), VII (F7), XI (F11), and XIII (F13B) were higher in the NMV/r group at day 15 (**Table S5**). The levels of complement component 3 (C3), aspartyl aminopeptidase (DNPEP), and C-type lectin domain family 13 member A (CD302) were higher in both the NMV/r and PBO/r groups at day 15 (**Figure 3D**). Enrichment analysis identified significant changes in modules related to myeloid cells/monocytes (M81), platelet and integrin activation (M85, M1.1), lysosomes (M209, M139), complement activation (M112.0, M112.1), and blood coagulation (M11.1) at day 15 in the NMV/r group compared to baseline (**Figure 3E,F**). Substantially higher monocyte-associated responses in COVID-19 infections have been associated with mild infection severity^21,22^. By week 10, the proteome returned to baseline states with no significant changes identified. Overall, a 15-day treatment of NMV/r induced transient changes in the plasma proteome that are known to modulate immune pathways associated with mounting effective antiviral responses, and these changes return to baseline levels by week 10.

**Figure 3.**
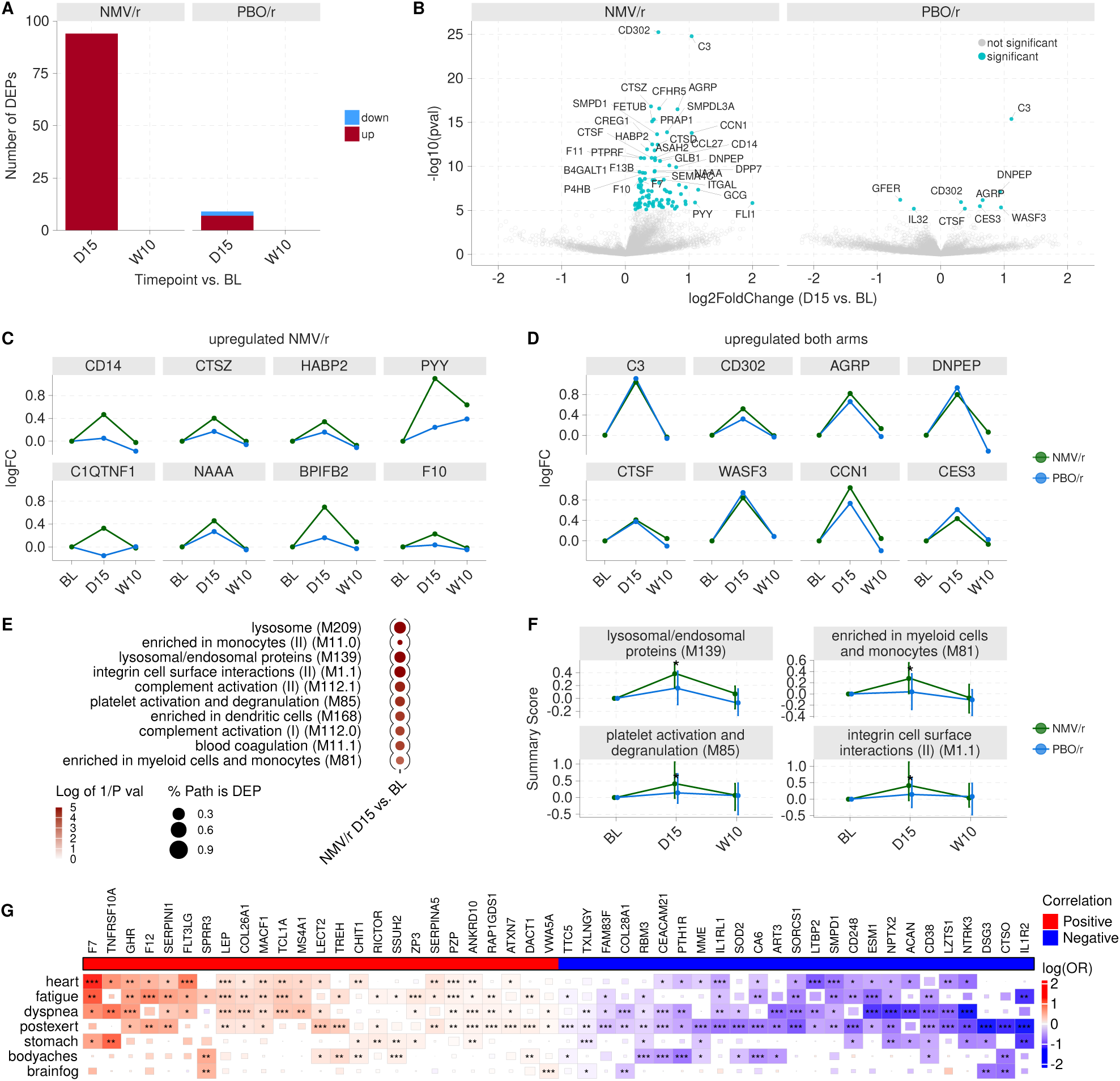
Plasma proteomic profiling during NMV/r treatment revealed transient immune pathway activation. (A) The number of upregulated (red) and downregulated (blue) differentially expressed proteins (DEPs) at each timepoint relative to baseline. (B) DEPs comparing day 15 versus baseline for the NMV/r group and PBO/r group. The y-axis is the unadjusted *P* values from within-group limma analysis adjusted for plate and accounting for repeated measures. Statistically significant proteins are colored blue with adjusted *P* <0.05 with Bonferroni correction. (C) Proteins with greater change from baseline in the NMV/r group. NMV/r (green), PBO/r (blue). (D) Proteins upregulated in both arms. (E) Blood immune pathways enriched at the day 15 timepoint compared with baseline for the NMV/r group. (F) Select pathways summary scores, normalized to an individual participant’s baseline. (G) Association between protein level and PRO severity in STOP-PASC at baseline as estimated by proportional odds logistic regression. The size of the squares and color is indicated by the log odds ratio (log(OR)) with significance indicated with asterisks: ***P≤0.001; **P≤0.01; *P≤0.05. The top 25 positively and negatively associated proteins with PRO severity ranked by p-value are shown.

In exploratory analysis, we sought to identify proteins that were associated with the severity of the core PROs reported by the participants in the STOP-PASC study. In total, 1327 proteins were associated with increasing or decreasing severity of one or more PROs (**Figure S8**), while 110 proteins demonstrated consistent patterns (p<0.05) with the trend present in three or more PROs (**Figure 3G**). For example, leptin (LEP), coagulation factor VII (F7), and coagulation factor XII (F12) were positively associated with fatigue and heart/cardiovascular symptoms; CD38 was negatively associated with dyspnea, and Interleukin 1 Receptor Like 1 (IL1RL1) and Interleukin 1 Receptor Type 2 (IL1R2) were negatively associated with post exertional malaise.

### Meta-analysis of Long COVID proteomics reveals conserved inflammatory signature

To expand beyond our STOP-PASC trial data and establish broader insights into LC proteomics, we turned to external publicly available LC cohorts. Multiple individual studies have reported differences in serum or plasma proteomes. However, there are currently no validated peripheral blood protein biomarkers for LC. Moreover, established biomarker signatures and endpoints to indicate treatment efficacy in LC clinical trials are lacking. To unify our understanding of the LC proteome, we curated publicly available Olink® data encompassing 590 total samples from 9 independent cohorts (**Table S6**)^8,9,11,12,23–27^. Next, we performed a comprehensive random-effects inverse-variance meta-analysis of 374 patients (n=52 Healthy Controls, n=76 Infected Recovered, n=246 LC) across six independent cohorts using MetaIntegrator (**Figure 4A**). We used three cohorts with 216 samples (n=22 Healthy Controls, n=61 Infected Recovered, n=133 LC) as validation cohorts. This approach has been successfully used for developing an FDA-cleared mRNA-based test for sepsis^28,29^. To enable more robust identification of LC-specific protein signatures that would remain regardless of the control population, we used two reference groups in the meta-analysis: LC vs. Healthy Controls (138 samples in three cohorts) or LC vs. Infected Recovered subjects (236 samples in three cohorts). Despite the biological and clinical heterogeneity present with LC and the technical heterogeneity across datasets, we identified 60 proteins that were differentially regulated in the same direction in independent cohorts (FDR<5%, effect size >0.5, proteins measured in ≥3 cohorts). We defined a LC Signature (LCS) score as the geometric mean of 60 upregulated proteins (**Figure 4B**). In the six discovery cohorts, the LCS score was significantly higher (p<0.05; **Figure 4C**) in patients with LC and distinguished them from Infected Recovered or Healthy Controls with a summary area under the receiver operating characteristic curve (AUROC) of 0.783 (95% CI: 0.578–0.988) (**Figure 4D**). In the three validation cohorts, the LCS score was significantly higher in LC patients compared to Healthy Controls or Infected Recovered patients (**Figure 4E**) with a summary AUROC of 0.759 (95% CI: 0.55-0.969) (**Figure 4F**). The LCS score was positively associated with older age; however, the LCS score did not exhibit significant sex-differences within patients with LC (**Figure S9**). Finally, longitudinal analysis of the STOP-PASC trial data revealed no significant treatment-associated changes in the LCS score from baseline to day 15 or baseline to week 10 between the NMV/r and PBO/r arms (**Figure 4G**). We also examined if any of the day 15 changes from baseline in LCS score differed between the patient-reported outcome trajectory classes we previously established in the STOP-PASC cohort^15^. Similar to our findings with other clinical endpoints, the LCS score changes did not differ between the participant trajectory classes (**Figure S10**). The results of this exploratory proteomic meta-signature endpoint are in line with the primary findings of the STOP-PASC trial showing no significant therapeutic benefit for improving select LC symptoms.

**Figure 4.**
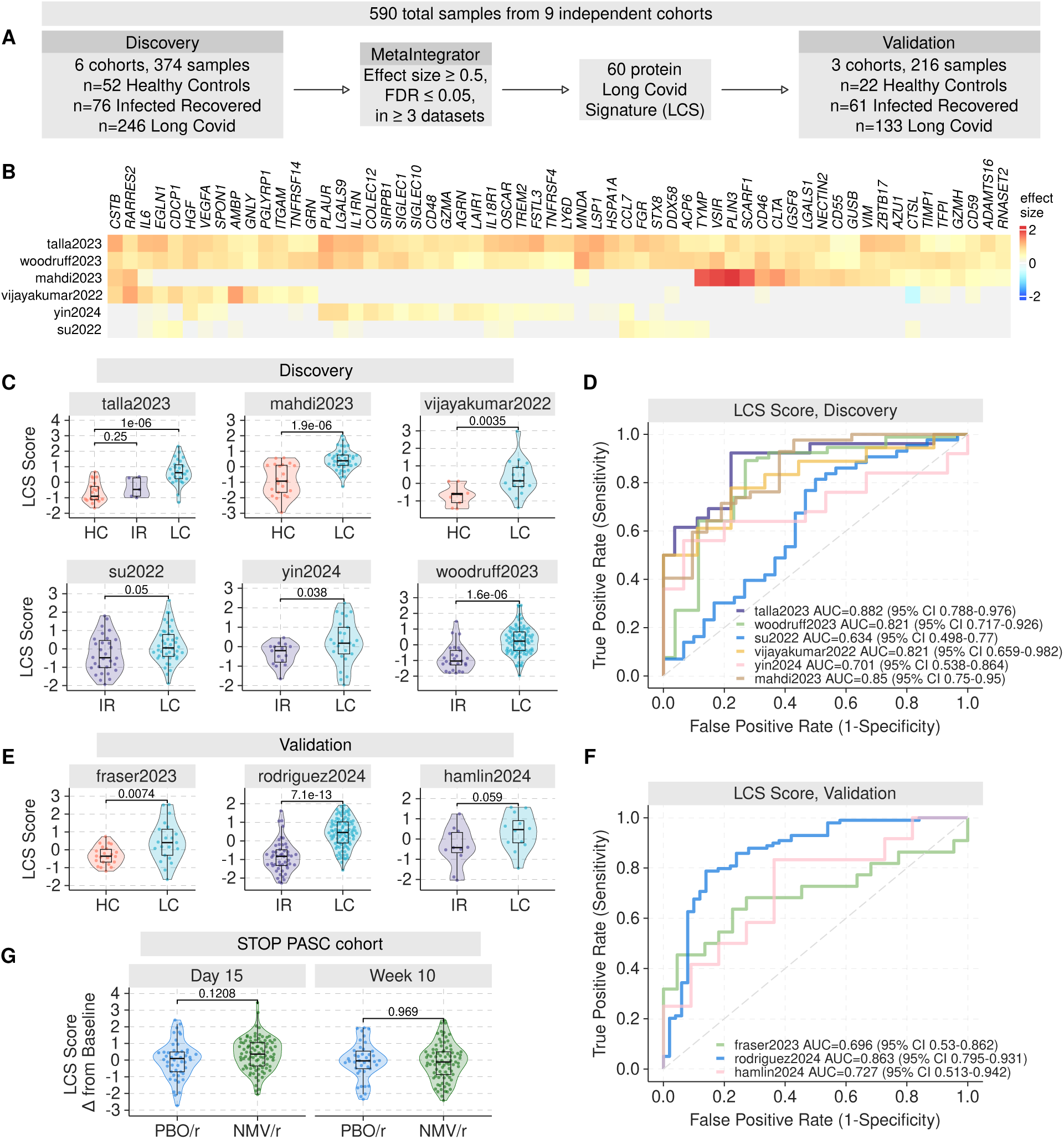
Multicohort meta-analysis identifies a conserved Long COVID proteomics signature. (A) Schematic of analysis pipeline. (B) Heatmap of proteins in the LCS score colored by effect size (Hedges’ g) comparing LC vs. Healthy Control or LC vs. Infected Recovered. Grey coloring indicates a protein that was not measured by the corresponding study which used an early version of Olink®. (C) Distribution of the LCS score in the discovery cohorts comparing LC vs. Infected Recovered or Long COVID vs. Healthy control (HC). Data are displayed as both violin plot and box and whisker plots. Statistical test: Wilcoxon rank-sum test. Healthy control (HC, red), Infected Recovered (IR, purple), Long COVID (LC, blue). (D) Receiver operating characteristic (ROC) curve in the discovery datasets distinguishing Long COVID vs. Healthy Control or Long COVID vs. Infected Recovered. (E) Distribution of the LCS score in validation cohorts. Statistical test: Wilcoxon rank-sum test. (F) ROC curve in the validation cohorts. (G) The change from baseline (day15-baseline and week10-baseline) of the LCS score comparing PBO/r and NMV/r groups. Statistical test: Wilcoxon rank-sum test.

### Long COVID involves sustained multi-compartment immune activation

To better understand the pathways, protein modules, and cell types associated with persistent immune dysregulation in LC, we applied a two-step approach to characterize the 60-protein LCS score. First, we performed a meta-analysis of proteomics pathways in LC using the six public discovery proteomics cohorts. We identified conserved upregulation of chemokines and receptors (M38) and myeloid/dendritic cell activation via the transcription factor NFkB (I) (M43.0) in LC (**Figure 5A**). Next, to map these protein-level signals to immune cell identities, we annotated the LCS score proteins by examining their corresponding RNA expression patterns in whole-blood single-cell transcriptomic data^30–33^ (n=584,260 cells, 223 samples) of healthy subjects and those with viral or bacterial infections. We adapted a methodology from our prior transcriptomic work, where we defined highly correlated gene expression sub-modules which are associated with increased or decreased risk of severe outcomes to acute respiratory viral infections^22^. Using this framework, we identified four primary immune cell-specific modules within the LCS score: a neutrophil module (including AZU1, PGLYRP1) a monocyte/dendritic cell module (including SIGLEC1/10, TREM2, FGR), a T/NK module (including GZMH, GZMA, GNLY), and a myeloid module with expression in both monocytes and neutrophils (**Figure 5B-D**). We then assessed each module in multiple independent LC proteomics cohorts, demonstrating each immune cell-specific module was elevated in LC patients compared to both healthy controls and infected-recovered individuals (**Figure 5E,F**). These findings support the premise that LC involves persistent activation across multiple immune compartments rather than dysregulation of a single cell type or pathway.

**Figure 5.**
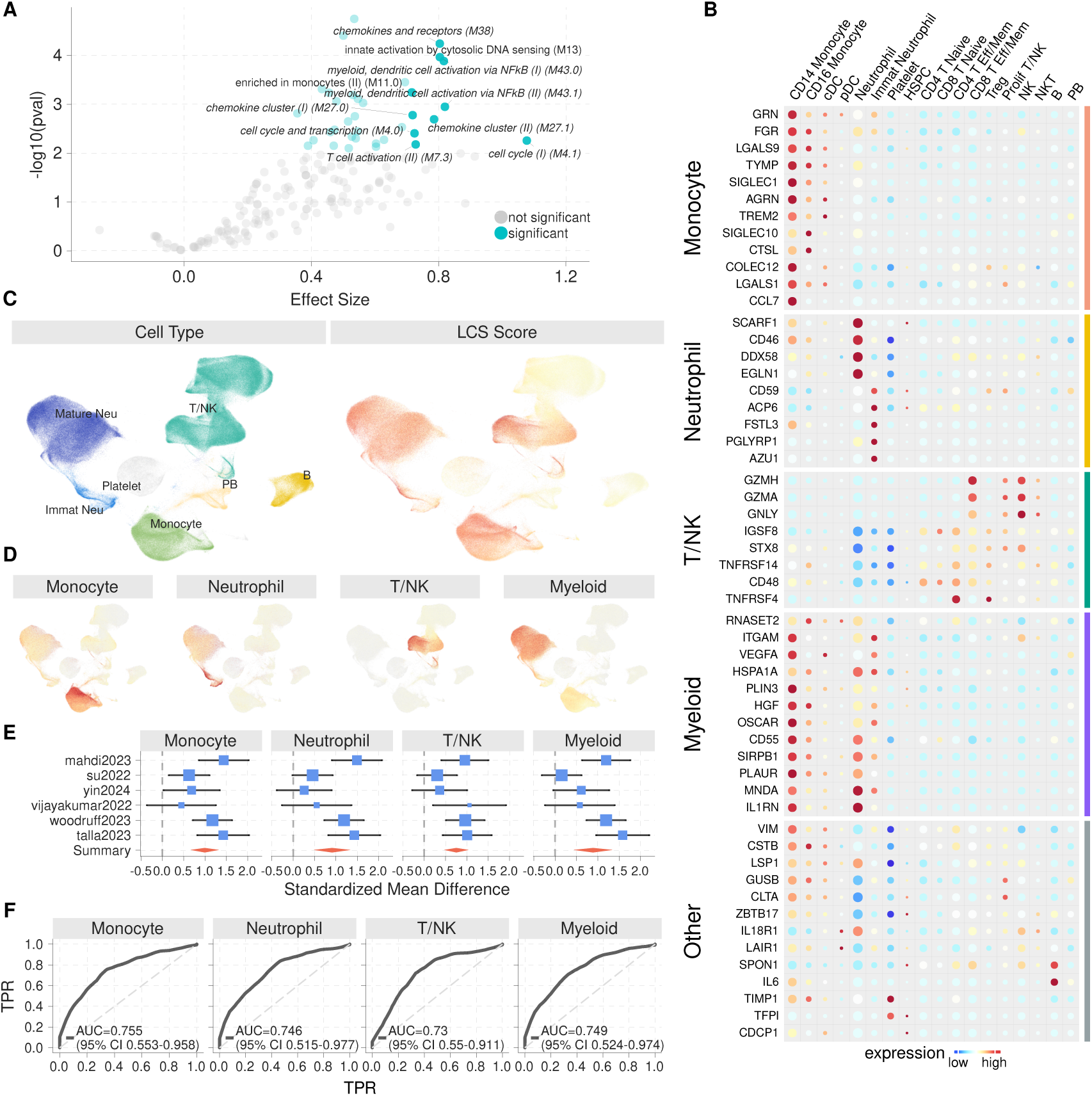
The LCS score is derived from multiple immune cell types with activation across neutrophil, monocyte, and T/NK cell compartments. (A) Proteomics meta-analysis of pathways enriched in Long COVID (n=6 studies). Pathways with with effect size FDR <0.05 are shown in blue, while non-statistically significant pathways are in grey. Labels indicate pathways with an effect size (Hedges’ g) above 0.7. (B) Heatmap of the average expression of the genes in single-cell whole-blood transcriptome data. (C) UMAP visualization of single-cell data colored by immune cell type and LCS Score. (D) Average score values for each immune module. (E) Proteomics meta-analysis of immune module scores upregulated in Long COVID. (F) Summary ROC curve for each immune module in the public proteomics discovery cohorts.

## Discussion

Long COVID research has yielded a complex landscape of immune system alterations and chronic inflammatory processes^7–9,25^. Here, we performed systems immunology profiling of the STOP-PASC trial cohort and analyzed responses to antiviral intervention. We identified significant immunological features that distinguish LC biology. Our proteomics findings revealed that nirmatrelvir-ritonavir (NMV/r) treatment induces transient changes in immune pathways, which are upregulated at day 15, but return to baseline levels by week 10. These results are in line with the clinical trial outcomes of STOP-PASC, where no sustained benefits for LC participants were observed in the treatment arm.

Nirmatrelvir is a selective inhibitor of the SARS-CoV-2 main protease (M^pro^) showing broad *in vitro* activity against human coronaviruses^34^. Inhibition of the SARS-CoV-2 M^pro^ renders it incapable of processing polyprotein precursors, preventing viral replication. In STOP-PASC LC participants receiving NMV/r, we observed upregulation of pathways linked to antiviral and pathogen clearance responses including myeloid and platelet activity, complement signaling, and lysosomal pathways and cathepsins, which are associated with phagocytic and antigen processing activity^7,35,36^. The proteomic changes we observed are consistent with nirmatrelvir’s established effectiveness for acute COVID-19^37^ and supports data that early antiviral treatment during acute COVID-19 may reduce LC risk^38–40^. Given the timing of intervention in our cohort which had protracted LC illness averaging more than 16 months, nirmatrelvir may not address the underlying immunological dysregulation in chronic LC. Alternatively, viral persistence may not be the primary driver of symptoms in our study participants. In line with our findings, the PAX LC clinical trial also showed no significant benefit in LC health outcomes for a 15-day NMV/r regimen^41^.

Ritonavir, used as a pharmacokinetic booster, inhibits CYP3A4 to slow nirmatrelvir metabolism and increase its systemic exposure, but has no direct antiviral activity against SARS-CoV-2^37^. One of the proteins upregulated at day 15 in both the NMV/r and PBO/r arms was complement component 3 (C3), which suggests this effect may be attributable to ritonavir rather than nirmatrelvir.

We detected only modest decreases in antibody levels against SARS-CoV-2 between baseline and week 10 in our STOP-PASC LC participants, suggesting relative stability in antibody levels over this timeframe. We observed a negative association between cardiovascular symptom severity and baseline antibody levels against several SARS-CoV-2 proteins, suggesting that more robust immune responses may be protective against certain LC manifestations. This is consistent with other reports which associated stronger immune responses, through higher levels of certain antibodies (including anti-RBD), as protective against cardiovascular LC symptoms^42^.

Our results suggest that measurement of PRO severity in LC aligns with meaningful underlying inflammatory signaling and biology. We observed leptin (LEP) was significantly associated with increased fatigue and several other PROs, consistent with reports that daily fatigue severity correlated significantly with serum leptin in women with chronic fatigue syndrome (CFS)^43^. This observation may indicate a shared mechanism in fatigue pathology across post-infectious syndromes including both CFS and LC, however, further research is needed to replicate this observation.

Previous evidence suggests reactivating latent herpesvirus may contribute to LC pathophysiology^6,9,44^. Our analysis did not identify differences in IgG antibody levels against EBV viral capsid antigen (VCA), nuclear antigen 1 (EBNA-1), early antigen (EA), or CMV glycoprotein B between STOP-PASC LC participants and individuals without LC. Our serological assessment at this later timepoint (protracted LC symptom duration) suggests that persistent herpesvirus reactivation was not a dominant feature in established LC, at least in our mostly vaccinated study population.

Even mild cases of acute SARS-CoV-2 infection can trigger the development of new (*de novo*) autoantibodies (AAbs) that target traditional autoantigens or cytokines^18,45^. While AAb repertoire changes return to homeostatic conditions after the acute phase in most patients, some with LC have detectable patterns of autoreactivity that could be associated with neurological sequela^45–47^. We did not observe widespread evidence of autoimmunity against traditional autoantigens in STOP-PASC LC participants. However, a subset of participants in our cohort showed reactivities against specific antigens (CENP A, fibrillarin, and PDC-E2). Epidemiological evidence and electronic health record studies continue to suggest increased risk of new-onset autoimmune conditions following SARS-CoV-2 infection^19,48–50^. However, our study was not powered to assess this as it would require longitudinal studies with thousands of participants. Similarly, our analysis of microclots found limited evidence of widespread coagulation abnormalities in this cohort. Still, this does not preclude the importance of endothelial dysfunction in specific LC subgroups who could benefit from targeted interventions^10^.

Here, we have identified a core set of 60 proteins displaying high effect sizes upregulated in LC, which can be further developed and translated into clinical assays. For example, further studies are needed to build a locked diagnostic machine learning classifier and to validate it on an independent, prospectively enrolled cohort. Despite considerable heterogeneity in LC, we have demonstrated via multicohort meta-analysis conserved dysregulation in inflammatory immune pathways. The proteins in the LCS Score which are consistently elevated in LC in multiple cohorts are associated with inflammation (IL6), monocyte (SIGLEC1/10) and neutrophil activation/granules (AZU1, PGLYRP1), and T/NK cell granzymes (GZMH, GZMA) suggesting a persistent dysregulated immune response. Furthermore, the multi-compartment immune inflammation we observed, involving neutrophil, monocyte, and T/NK cell modules, suggests ongoing immune activation that persists beyond the acute infection phase. This persistent immune activation could contribute to the diverse symptomatology observed in LC patients and may explain why targeted approaches addressing single pathways have shown limited efficacy. Finally, the conserved inflammatory signature derived here may serve as a candidate biomarker endpoint or as a predictive enrichment tool to stratify patients in future LC clinical trials, helping to guide targeted therapeutic evaluation.

## Limitations

Our study has several limitations. First, LC consists of significant individual heterogeneity, making it difficult to identify universally applicable biomarkers and mechanisms across diverse patient populations. The robustness of our immune-symptom associations may be limited by our small cohort size and patient diversity. Our study only focused on circulating biomarkers measurable in the peripheral blood and not tissues, which can involve organ-specific dysfunction which may not be adequately reflected in blood biomarkers. Additionally, we measured most biomarkers at specific timepoints (baseline, day15, week10), which may not capture dynamic temporal changes throughout the fluctuating LC disease course. Due to the 2:1 randomization study design that allocated more participants to the NMV/r arm than to the PBO/r arm, our study had greater statistical power to detect proteomics changes in the NMV/r subgroup compared to the PBO/r subgroup. Lastly, while certain immune features correlated with symptom burden, our study cannot infer whether these features drive patient-reported outcomes or result from them. Future clinical trials are needed to evaluate effective interventions and biomarkers of treatment response for this complex and disabling condition.

## Methods

### Study Population

Baseline demographics and full eligibility criteria of the STOP-PASC cohort have previously been reported^13^. Briefly, the STOP-PASC cohort enrolled adults (≥18 years) with clinician determined Long COVID (LC) symptoms persisting more than 90 days after their initial COVID-19 infection and presenting with at least 2 moderate or severe core symptoms or symptom clusters defined as fatigue, brain fog, body aches, cardiovascular symptoms, shortness of breath (dyspnea), and gastrointestinal symptoms. LC participants in the STOP-PASC study (n=155) were randomized 2:1 to receive either oral nirmatrelvir-ritonavir (NMV/r, n=102) or placebo- ritonavir (PBO/r, n=53) for 15 days. Symptom surveys, patient-reported outcomes, and biological samples were collected from participants at baseline and four follow-up time points until last visit at 15 weeks. This systems immunology profiling utilized n=152 participants (n=100 NMV/r, n=52 PBO/r) with sufficient post-baseline biological samples. This study was approved by the Stanford Institutional Review Board. All participants provided written consent.

For comparative analyses against the STOP-PASC LC participants, two control cohorts were also included in the immune profiling. The LC negative (LC^neg^) population consisted of 42 individuals without LC, comprising 30 Pfizer employees and 12 samples from the Institute of Systems Biology (ISB). Inclusion criteria for the LC^neg^ group were: 1) no COVID-19 infection within 90 days, 2) no vaccination within 28 days, and 3) no Long COVID symptoms. Additionally, 42 healthy uninfected pre-pandemic control samples were obtained from ISB. Autoantibody profiles and viral serological responses were compared between STOP-PASC participants, LC^neg^ individuals, and pre-pandemic controls.

### Biospecimen collection, processing, and plate design

We report systems immunology profiling analysis of EDTA plasma collected from the STOP-PASC participants. We previously reported analysis of nasal swabs, stool specimens, and digital biometric measures^13,14^. We used a combination of Enzyme-Linked Immunosorbent Assay (ELISA) and multiplexed assays to assess immune profiles including plasma protein profiling with Olink® Explore HT (n=5400 analytes) using blood samples collected at day 0, day 15, and week 10; IgG plasma autoantibodies using a 76-plex microbead-based protein array at day 0 and week 10; and anti-viral IgG responses using plasma collected at day 0 and week 10, using a combination of microbead arrays and multiplexed chemiluminescence measurements (Meso Scale Discovery, MSD); and assays for microclots. To minimize freeze/thaw cycles, master plates were prepared and distributed to collaborators. These plates were balanced for demographic factors (age, sex, race, and ethnicity) and designed to ensure that longitudinal samples from the same patient were analyzed on the same plate.

### Plasma protein profiling using Olink® Explorer HT NGS

Plasma proteomics was analyzed using Olink® multiplex proximity extension assay (PEA) with the Olink® Explore HT 5400 panel (Olink proteomics; www.olink.com) according to the manufacturer’s instructions. The PEA is a dual-recognition immunoassay in which two matched antibodies labeled with unique DNA oligonucleotides simultaneously bind to a target protein in solution. This proximity binding allows their DNA oligonucleotides to hybridize, serving as a template for a DNA polymerase-dependent extension step. This process creates a double-stranded DNA "barcode" that is unique for the specific antigen and quantitatively proportional to the initial concentration of the target protein. Following hybridization and extension, the reaction proceeds to PCR amplification, and the resulting amplicon is quantified by next generation sequencing on an Illumina NovaSeqX Plus. Olink® Explore HT software version 6.7.2 was used for quality control. Normalized Protein expression (NPX) values were used for downstream analysis.

### Differential protein expression analysis

We performed differential protein analysis of the STOP-PASC trial Olink® proteomics data using the *limma* R package to calculate fold changes^51^. For within-group analysis of differential proteins, we used linear mixed-effects models adjusting for plate as a fixed effect and accounting for repeated measures on the same participants across multiple timepoints. Subject IDs were set as random effects in the block argument. We estimated the correlation between repeated measurements using the *duplicateCorrelation* function in *limma.* We ensured longitudinal samples from the same participant (baseline, day 15, week 10) were run on the same plate. Plate designs were randomized/balanced for age, sex, ethnicity, and race prior to sample processing. The *P* values from *limma* modeling were adjusted by Bonferroni correction. A small number of participants had new infections during the trial (at some time between D15 and W10) based on spike and nucleocapsid antibodies. Sensitivity analysis was performed by removing these participants in each group (NMV/r n=5, PBO/r n=3) when comparing W10 vs. baseline and no differences were observed.

### Protein set overrepresentation enrichment analysis

Protein-based over-representation analysis (ORA) was performed on differentially expressed proteins (adjusted *P*<0.05) identified from the NMV/r day 15 vs. baseline comparison. We calculated the enrichment score of immune modules from The Blood Transcriptional Modules (BTM) gene sets^52^ which were translated to the protein level. The p-values were adjusted using Bonferroni correction. Next, for longitudinal analysis, we summarized the protein pattern of each immune pathway by computing a summary score as the geometric mean of the NPX of all proteins in the pathway, normalizing to each patient’s baseline.

### Electrochemiluminescence Assays for Plasma IgG Antibodies

Plasma samples from treatment and placebo group participants were heat inactivated at 56°C for 30 minutes then assayed for IgG antibodies using multiplexed electrochemiluminescence assays in 96-well plate format with MSD® V-PLEX® serology panels and instrumentation according to the manufacturer’s instructions. The V-PLEX COVID-19 Coronavirus Panel 2 kits (Meso Scale Discovery Cat#K15369U) was used to detect IgG specific for SARS-CoV-2 Nucleocapsid, Spike, S1 N-terminal domain (NTD) and receptor binding domain (RBD), as well as for Spike proteins of SARS-CoV-1 and other HCoVs including HCoV-OC43, HCoV-HKU1, HCoV-NL63, and HCoV-229E. The V-PLEX Respiratory Panel 1 (IgG) Kit (Meso Scale Discovery Cat#K15365U) was used to measure IgG specific for Influenza hemagglutinins A/Hong Kong/2014 H3, Flu A/Michigan/2015 H1, Flu A/Shanghai/2013 H7, Flu B/Brisbane/2008 HA, Flu B/Phuket/2013 HA as well as Respiratory Syncytial Virus (RSV) Pre-Fusion F. The V-PLEX SARS-CoV-2 Panel 27 (IgG) Kit (Meso Scale Discovery Cat#K15606U) was used to assay the IgG concentration to SARS-CoV-2 Spike, SARS-CoV-2 Spike (B.1.351), SARS-CoV-2 Spike (B.1.617.2; AY.4), SARS-CoV-2 Spike (BA.2), SARS-CoV-2 Spike (BA.2.12.1), SARS-CoV-2 Spike (BA.2+L452M), SARS-CoV- 2 Spike (BA.2+L452R), SARS-CoV-2 Spike (BA.3), SARS-CoV-2 Spike (BA.4), SARS-CoV-2 Spike (BA.5). Plasma samples were analyzed in duplicate at a 1:5,000 dilution in MSD Diluent 100, and a 7-point calibration curve of MSD Reference Standard 1 and MSD Serology Control 1.1, 1.2, and 1.3 was also measured. The electrochemiluminescence detection agent was Sulfo-tag conjugated anti-human IgG (Meso Scale Discovery Cat#D21ADF-3).

### Microclot measurement

Thioflavin T fluorescent staining was utilized to measure fibrin amyloid microclots in plasma, using a protocol adapted from Pretorius et al.^20,53^. Briefly, plasma samples were incubated with 5µM Thioflavin T for 30min at RT, protected from light. Following incubation, samples were diluted 10-fold with HBSA (20mM HEPES, 150mM NaCl, 0.1% bovine serum albumin, pH 7.4) in a black 96-well plate. Total fluorescence at the excitation wavelength of 450±10nm and the emission at 482±10nm was measured using a BioTek Cytation 5 fluorescence plate reader with Gen5 software (Agilent Technologies, Santa Clara, CA, USA). Results were normalized to the fluorescence value of fibrinogen-immunodepleted plasma, as a negative control. All samples were measured in triplicate, and measurement was repeated if the variation between replicates exceeded 15%. These results are reported as a ratio of the read against the corresponding plate’s negative control (averaged across the three negative control reads in that plate). We report the results from averaging across each person’s three reads, where a higher value corresponds to more clotting, with values above 2 considered evidence for microclotting. Standardized mean differences (SMD) are reported to assesses the magnitude of differences: a larger SMD indicates a larger difference between the groups (e.g. 0.2 = small difference, 0.5 = moderate difference, 0.8 = large difference).

### Bead-based antigen arrays

We created two different custom, bead-based antigen arrays. The “COVID-19 Viral Array” included recombinant, purified SARS-CoV-2 proteins from commercial sources, or recombinant proteins produced in the labs of Peter Kim and Taia Wang (**Table S2**). We also included proteins or protein fragments from SARS-CoV- 1, Middle East Respiratory Syndrome (MERS), seasonal coronaviruses (OC43, 229E, NL63, and HKU1), Hepatitis B, Mumps, Rubella, Rubeola, Ebola, Epstein-Barr Virus, and Influenza (hemagglutinin (HA) from A/California/07/2009 H1N1). The cytokine/traditional autoantigen array was composed of cytokines, chemokines, growth factors, acute phase proteins, cell surface proteins and commercial protein antigens associated with connective tissue diseases (**Table S3**). Each array was constructed and used for probing as previously described with modifications^18^.

In short, antigens were conjugated to uniquely barcoded carboxylated magnetic beads (MagPlex-C, Luminex Corp.). For each assay, the bead array was distributed into a 384-well plate (Greiner Bio-One) by transfer of 5 µL bead array per well. In total, 45 µL of diluted plasma sample was transferred into the 384-well plate containing the bead array. Samples were incubated for 60 minutes on a shaker at room temperature. Beads were washed with 3 × 60 µL PBS-Tween on a plate washer (EL406, Biotek) and then incubated with 50 µL of 1:1,000 diluted R-phycoerythrin–conjugated (R-PE–conjugated) Fc-γ–specific goat anti–human IgG F(ab’)2 fragment (Jackson ImmunoResearch, 106-116-098) for 30 minutes. The plate was washed with 3 × 60 µL PBS-Tween and resuspended in 50 µL PBS-Tween prior to analysis using a FlexMap3D instrument (Luminex Corp.). Prototype human plasma samples derived from participants with AIDs with known reactivity patterns (e.g., Scl-70, centromere, SSA [Ro], SSB [La], whole histones, RNP, anti-IFN) were used as positive controls. Binding events were displayed as median fluorescence intensity (MFI). For normalization, MFI values for unconjugated, bare bead IDs were subtracted from MFI values for each antigen-conjugated bead ID for each sample.

Plasma samples were considered “positive” for antibodies recognizing a specific antigen if the normalized MFI was > 5 standard deviations (SD) above the average MFI for HC for that antigen, and if the normalized MFI was >3,000 units, a more stringent threshold than those commonly published in related literature^18^.

### Associations of viral serology and protein levels with patient-reported outcomes (PROs)

This analysis involved testing the associations between immune profiles (viral serology or protein levels) and clinical features. Proportional odds logistic regression models were fit for ordinal severity level of each core symptom at baseline. The core symptoms were based on a Likert scale score: fatigue, brain fog, dyspnea, body aches, gastrointestinal symptoms, and cardiovascular symptoms. Each model was adjusted for the batch plate in which the sample was run.

### Meta-analysis of Long COVID Proteomics

We used the R package *MetaIntegrator* to identify differentially expressed proteins between Long COVID and Healthy Patients or Long COVID and Infected Recovered patients as described previously^54^. First, we computed the effect size (Hedges’ adjusted g) for each protein in each study. We combined effect sizes across all studies for each protein using a random-effects inverse variance model (DerSimonian-Laird method^55^), which weights the effect size by the inverse of the variance in that study. We correct p-values for the summary effect sizes of each protein for multiple hypothesis testing with the Benjamani-Hochberg correction to control the False Discovery Rate (FDR). Briefly, we downloaded Olink Proteomics data from 9 independent studies with 590 samples. Study details for all datasets are listed in **Table S6**. We limited analyses to serum or plasma proteomics from blood. The six cohorts used in discovery were required to have both Long COVID patients and Healthy or Infected Recovered for comparison. While inclusion criteria for Long COVID varied slightly by study, all defined Long COVID as symptoms persisting beyond the acute infection phase (ranging from 2-3 months to 18 months). Infected Recovered are individuals with prior SARS-CoV-2 infection but without persistent symptoms. We used the following criteria to select proteins in the Long COVID Signature (LCS) Score: Effect Size>0.5, FDR<0.05, and proteins which were measured in at least three of the discovery cohorts. Next, we define the LCS Score of a sample as the geometric mean of the 60 over-expressed proteins. The geometric means across samples in the same study are scaled with a z-score. We used the Mann–Whitney U test (Wilcoxon rank-sum test) to compare LCS scores between two groups. When calculating the change from baseline (week10-baseline) of the LCS score in the STOP-PASC trial comparing PBO/r and NMV/r groups, we normalized to each participant’s baseline. Sex-differences in the LCS score were assessed via Wilcoxon rank-sum test. We examined the relationship between age and the LCS score by calculating Spearman’s rank correlation coefficient.

We also performed a meta-analysis of Long COVID proteomics pathways. First, we summarized each immune pathway by computing the geometric mean of the NPX of all proteins in the pathway. Next, we compute the summary effect sizes and combine with random-effects inverse variance model as before. Pathways meeting the significance threshold (FDR < 0.05) were considered differentially regulated in Long COVID patients.

### Calculation of the LCS Score

The LCS score is computed as the geometric mean of the following 60 proteins: {PLAUR, CSTB, RARRES2, IGSF8, VIM, LGALS1, ZBTB17, CCL7, SIRPB1, CDCP1, COLEC12, PGLYRP1, NECTIN2, SPON1, CD46, GRN, SIGLEC1, ITGAM, FGR, SIGLEC10, GZMH, TIMP1, CD48, CLTA, CTSL, CD59, GZMA, EGLN1, TREM2, LGALS9, ADAMTS16, ACP6, TFPI, VEGFA, AGRN, AZU1, RNASET2, GNLY, IL18R1, TNFRSF14, DDX58, GUSB, STX8, LAIR1, OSCAR, TNFRSF4, VSIR, IL6, MNDA, SCARF1, TYMP, CD55, HGF, LY6D, AMBP, PLIN3, HSPA1A, IL1RN, LSP1, FSTL3}.

### ROC curves

The summary area under the receiver operating characteristic curve (AUROC) represents a weighted average of multiple independent ROC curves. A perfect classifier will have an AUROC of 1, while a random classifier will have an AUROC of 0.5. True positive rate (TPR) values for each curve were approximated using linear interpolation, and the summary ROC curve was calculated using the mean of the TPR values for each curve, weighted based on sample size. A weighted standard deviation was also calculated for each TPR. The AUROC was calculated using the trapezoid rule. We computed 95% CIs for individual AUCs using the DeLong method as implemented in the *pROC* package in R^56,57^.

### Protein modules

For each protein in the LCS Score, we used single-cell transcriptomic data (scRNA-seq) to calculate the percentage of cells within each cell type that expressed the corresponding gene. We analyzed peripheral blood scRNA-seq data (584,260 cells, 223 samples) from four studies of patients with viral or bacterial infections and healthy controls, which was integrated in Seurat and cell types were annotated as described previously^58^. To define the cell-type specific modules we compared the expression between the cell population of interest against other immune cells in peripheral blood. We used the following criteria for gene signature selection: 1) the gene is expressed in more than 50% of cells in the cell population of interest and less than 20% of all other cell types expressing it or 2) the gene has an average log2 fold change ≥ 1.5 in the cell population of interest compared to all other immune cells. Genes not meeting these criteria were labeled as myeloid (e.g., both neutrophil/monocyte) or other.

### Statistical Analysis

All statistical tests were two-sided, and results with p<0.05 were considered statistically significant. For comparisons between two independent groups (in antibody levels, proteomics measurements, etc.), we used the Mann-Whitney U test (Wilcoxon rank-sum test). We corrected p-values for multiple hypotheses testing using the Benjamini-Hochberg correction to obtain the false discovery rate (FDR) unless otherwise specified. For analyses requiring adjustment for multiple comparisons, significant results were determined after FDR correction with the same significance threshold (FDR<0.05). All analysis was performed in R v4.3.2.

## Figures

Figures were generated using *ggplot2* and *ComplexHeatmap*^59^. Illustrations were used from NIAID NIH BioArt Source (bioart.niaid.nih.gov/bioart/) #52, #250, #464, and #501.

## Author Contributions

*<u>Conceptualization:</u>* Geng, Utz, Khatri, Kwon, Prestwood, Singh, Maestri

*<u>Analysis Design and Methodology:</u>* Maestri, Khatri, Utz, Prestwood, Geng, Liang

*<u>Analysis or interpretation of data:</u>* Maestri and Zheng analyzed Olink data. Maestri, Kwon, and Prestwood analyzed autoantibody data and viral serology data. Maestri and Liang analyzed microclot data. Maestri, Zheng, and McCann analyzed single-cell data.

*<u>Drafting of the manuscript</u>:* Maestri, Utz, Geng, Khatri

*<u>Critical review of the manuscript:</u>* Maestri, Khatri, Utz, Geng, Bonilla, Jagannathan, Hedlin, Liang, Singh, Wood, Heath, Guthridge, Kwon, Prestwood, Boyd

*<u>Statistical analysis:</u>* Maestri, Zheng, Hedlin, Liang, McCann, Shaw, Tian, Khatri

*<u>Data collection/acquisition and experimental design:</u>* Kwon and Prestwood processed biospecimen, performed experiments, generated autoantibody and Luminex viral serology data. Jones, Lu, Wiley, and Guthridge generated Olink data. Haraguchi, Wirz, Afaghani, Lam, and Boyd generated MSD viral serology data. D Mahmood, Phillips, Sim, and Wood generated microclot data. Heath contributed data and resources.

*<u>Obtained funding:</u>* Geng, Utz, Singh

*Administrative, technical, or material support:* Utz, Kwon, Prestwood, Singh, Geng

*<u>Supervision:</u>* Khatri, Utz, Geng, Jagannathan, Singh

## Funding

This study was funded by Pfizer Inc. Maestri is funded by the Ford Predoctoral Fellowship and NIH T32AI007290. Utz was funded by Pfizer and the Stanford Department of Medicine Team Science Program. Khatri is funded by Bill and Melinda Gates Foundation (OPP1113682, INV-076306, INV-070250), the National Institute of Allergy and Infectious Diseases (NIAID) grants U19AI057229 and U19AI67903, NIAID contract 75N93022C00052, and the Brennan Family. The Stanford Quantitative Sciences Unit was partially supported by the NIH (grant UL1 TR003142). The Stanford REDCap platform, which was used to conduct the study’s prescreening survey was developed and is operated by the Stanford Medicine Research IT team and is subsidized by the Stanford School of Medicine Research Office, and the NIH’s National Center for Research Resources and the National Center for Advancing Translational Sciences (UL1 TR001085). The Stanford Clinical and Translational Research Unit is supported by the Stanford CTSA Award from the NIH’s National Center for Advancing Translational Science (NIH-NCATS-CTSA grant 5UL1TR003142).

## Role of the Funder/Sponsor

The funder had no role in the conduct of the study; collection, management, and analysis of the data; preparation of the manuscript; and decision to submit the manuscript for publication.

## Acknowledgements

We thank the members of the Khatri Lab and Utz lab for their insights and helpful discussion. We thank Petter Brodin, Lucie Rodriguez, Nadia Roan, Timothy Henrich, and James Heath for sharing data with us. We thank Ashley Utz and the Peter Kim Lab for providing antigens and other reagents for this study.

## Declaration of Interests

Dr Utz reported receiving grants from Pfizer during the conduct of the study. Dr Khatri reported receiving grants from Pfizer and grants from Eli Lilly, and is co-founder and consultant to Inflammatix, Inc. Dr Hedlin reported receiving institutional support from Pfizer and grants from National Institutes of Health (NIH) outside the submitted work. Dr Bonilla reported receiving grants from Pfizer during the conduct of the study. Dr Jagannathan reported receiving grants from Pfizer during the conduct of the study. Dr Singh reported receiving grants from Pfizer during the conduct of the study. Dr Geng reported receiving grants from Pfizer during the conduct of the study; grants from Agency for Healthcare Research and Quality and NIH outside the submitted work; and personal fees from Clearview Healthcare Partners and Speakers Network outside the submitted work. Dr Boyd reported receiving grants from Pfizer during the conduct of the study, has consulted for Regeneron, Sanofi, Novartis, Genentech, Visterra and Janssen on topics unrelated to this study, owns stock in AbCellera Biologics, and is a co-founder of Immunera Inc. Dr Wood reported receiving grants from Pfizer on topics unrelated to this study. The other authors declare no competing interests.

## Data and code availability

All software utilized in this study are publicly available. Code used to generate figures will be available on Github (https://github.com/Khatri-Lab/STOP_PASC_biomarkers) upon publication. Generalized code for Olink HT data quality control can be found on Github (https://github.com/guthridge-informatics/Olink_HT). The Olink data, autoantibody and viral arrays, and MSD serology generated in this study will be available upon publication. Any additional information required to reanalyze the data reported in this paper is available from the lead contact upon request.

---

**Figure S1.**
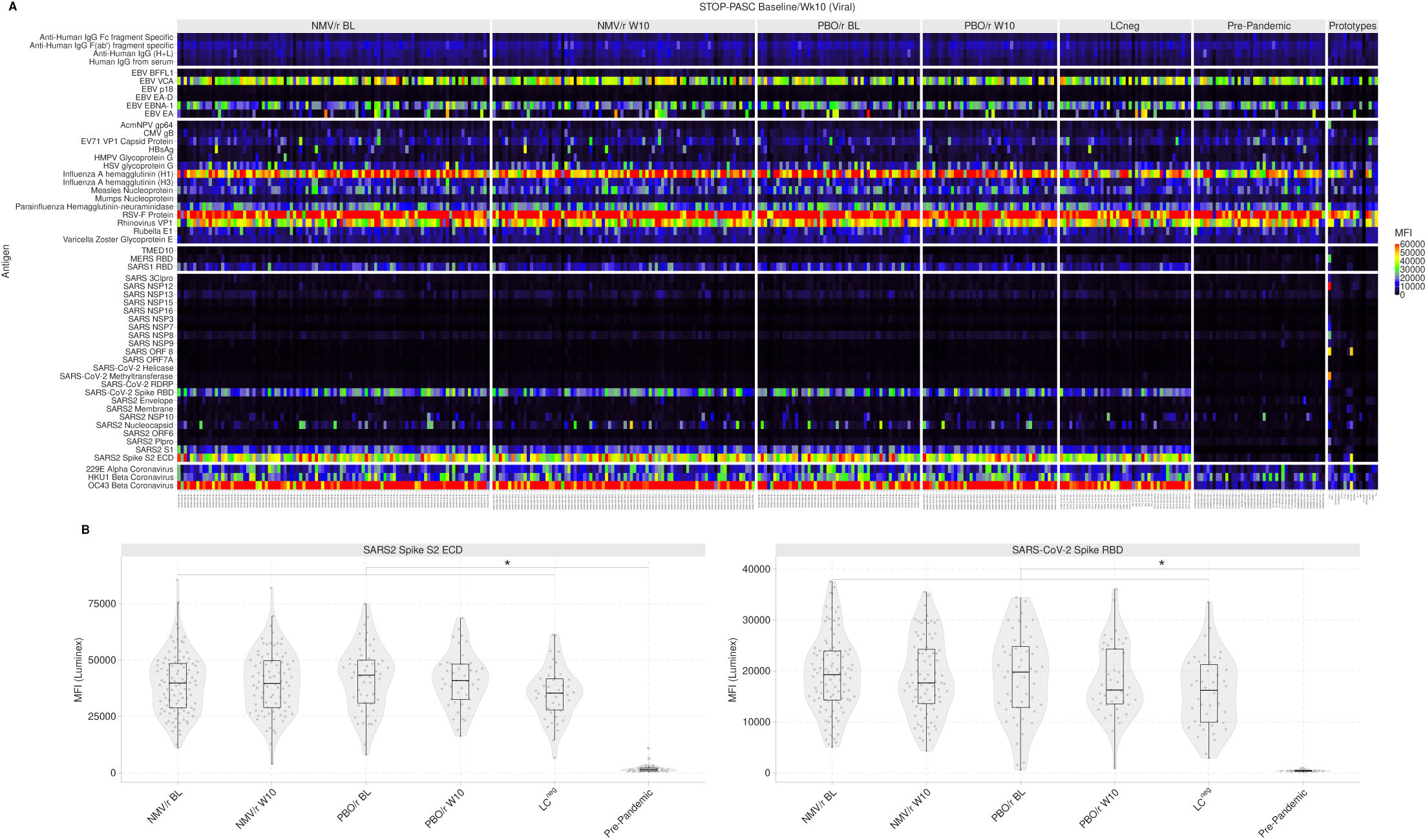
Viral Luminex panel. (A) STOP-PASC IgG serology using a panel of various SARS-CoV-2 proteins, herpesviruses, and seasonal coronaviruses. (B) SARS-CoV-2 Spike RBD and Spike S2 ECD are only positive during pandemic-era samples.

**Figure S2.**
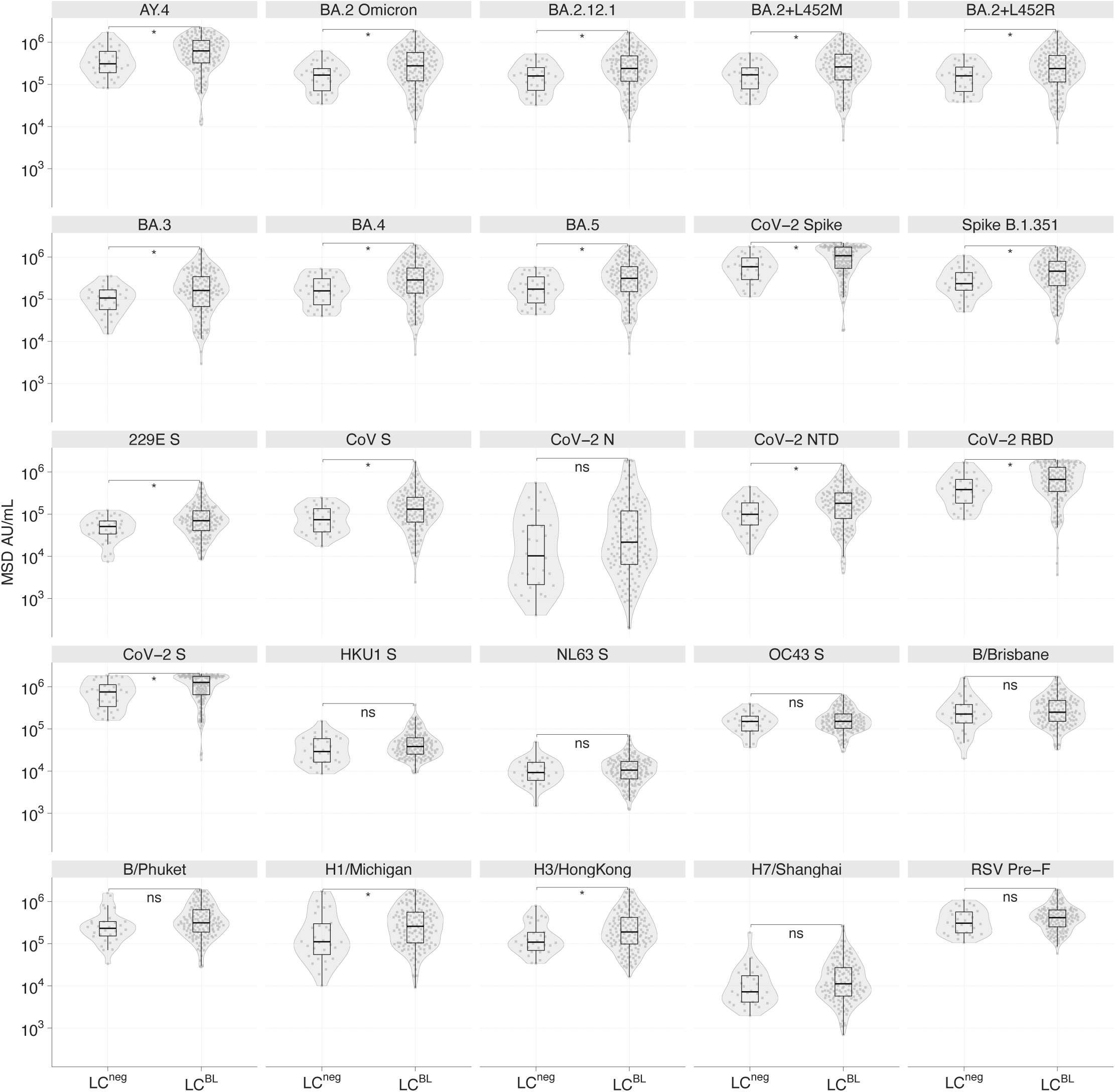
Magnitude of anti-SARS-CoV-2 IgG antibody levels in MSD arbitrary units (AU/mL). Statistical test: Wilcoxon rank-sum test with FDR correction (Benjamini-Hochberg).

**Figure S3.**
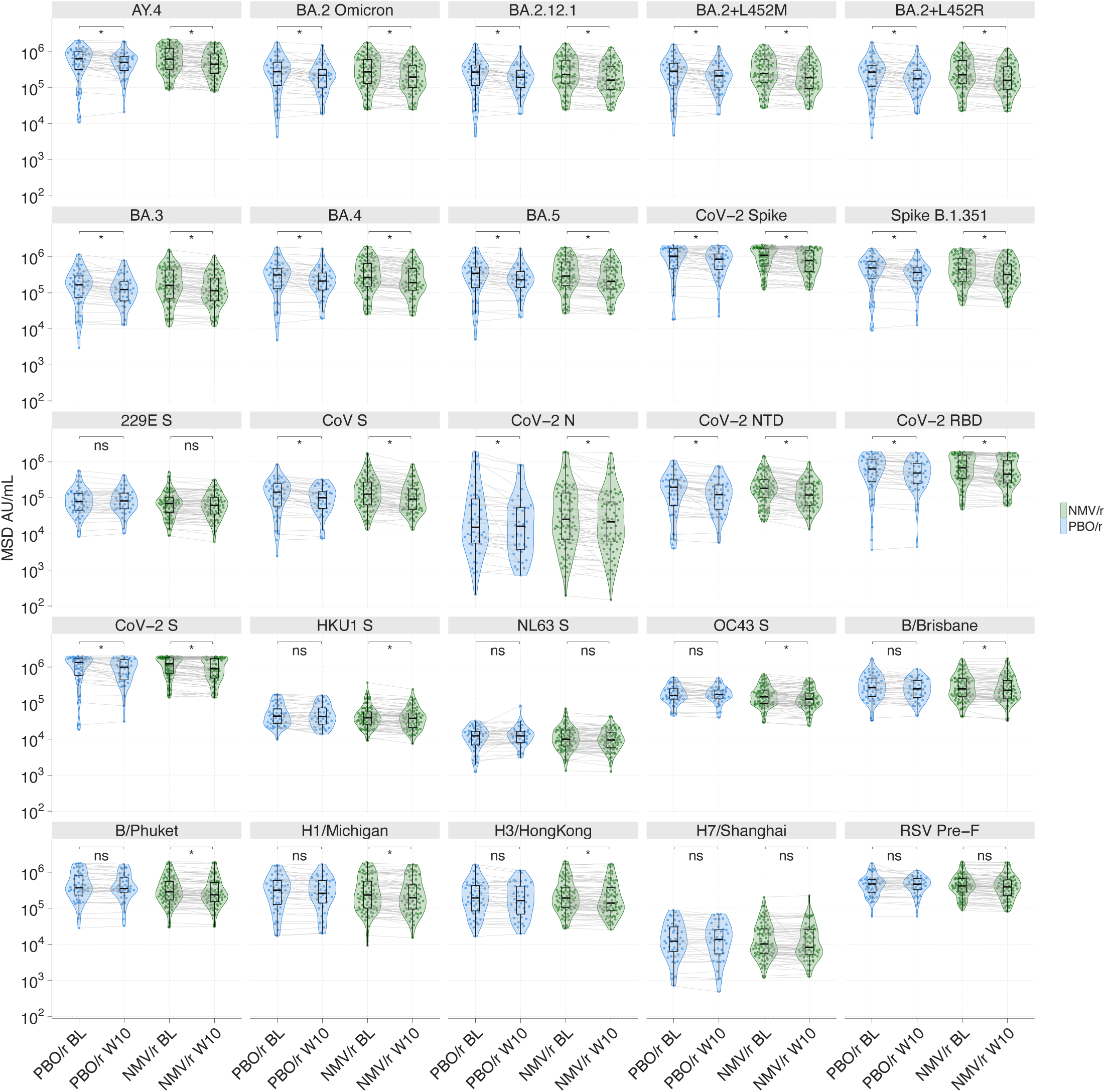
Magnitude of anti-SARS-CoV-2 IgG antibody levels in MSD arbitrary units (AU/mL) between baseline and week10 in NMV/r (green) and PBO/r arms (blue). Statistical test: Paired Wilcoxon rank-sum test with Bonferroni correction.

**Figure S4.**
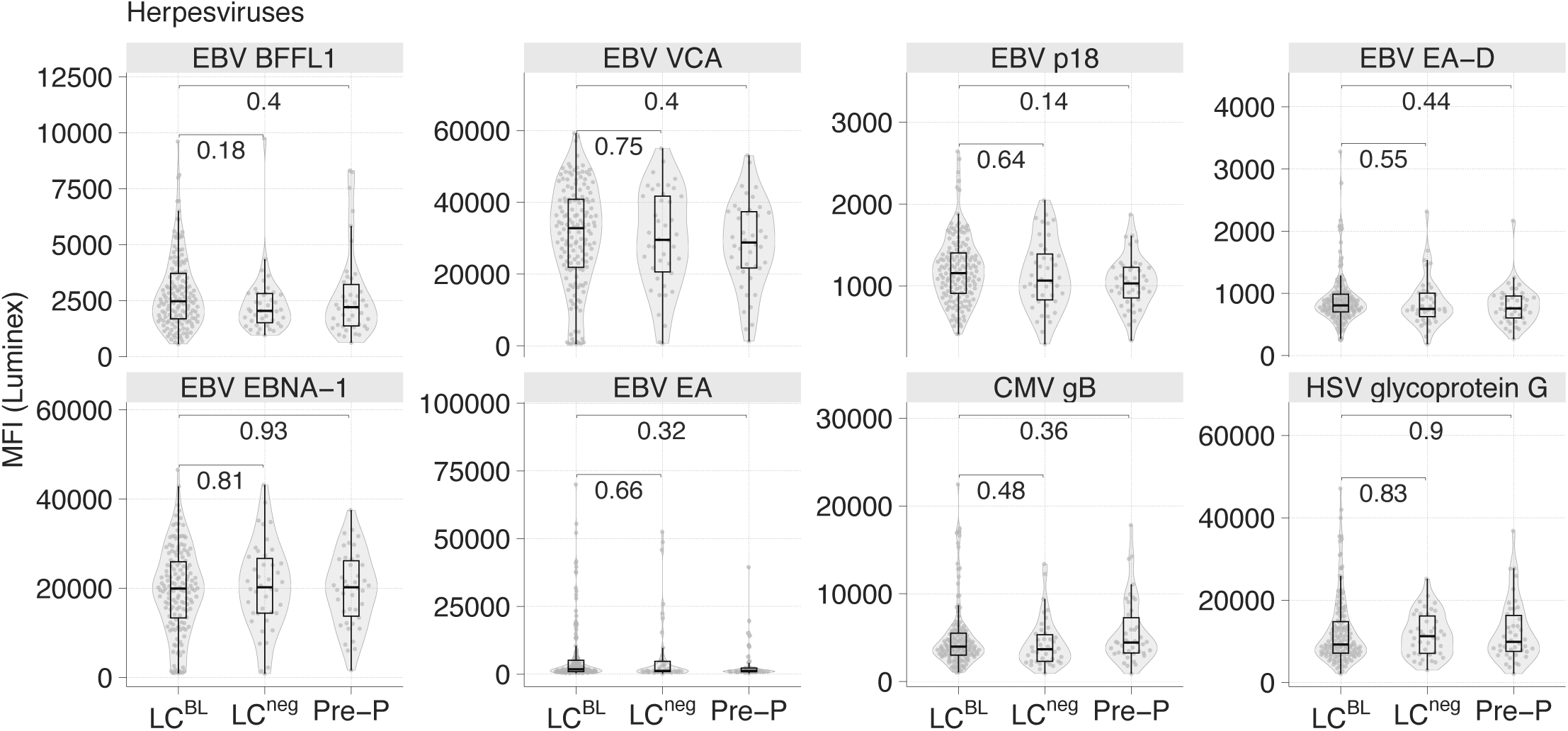
Herpesvirus antibody levels. Luminex results for EBV, CMV, and HSV. Statistical test: Wilcoxon rank-sum test with FDR correction (Benjamini-Hochberg).

**Figure S5.**
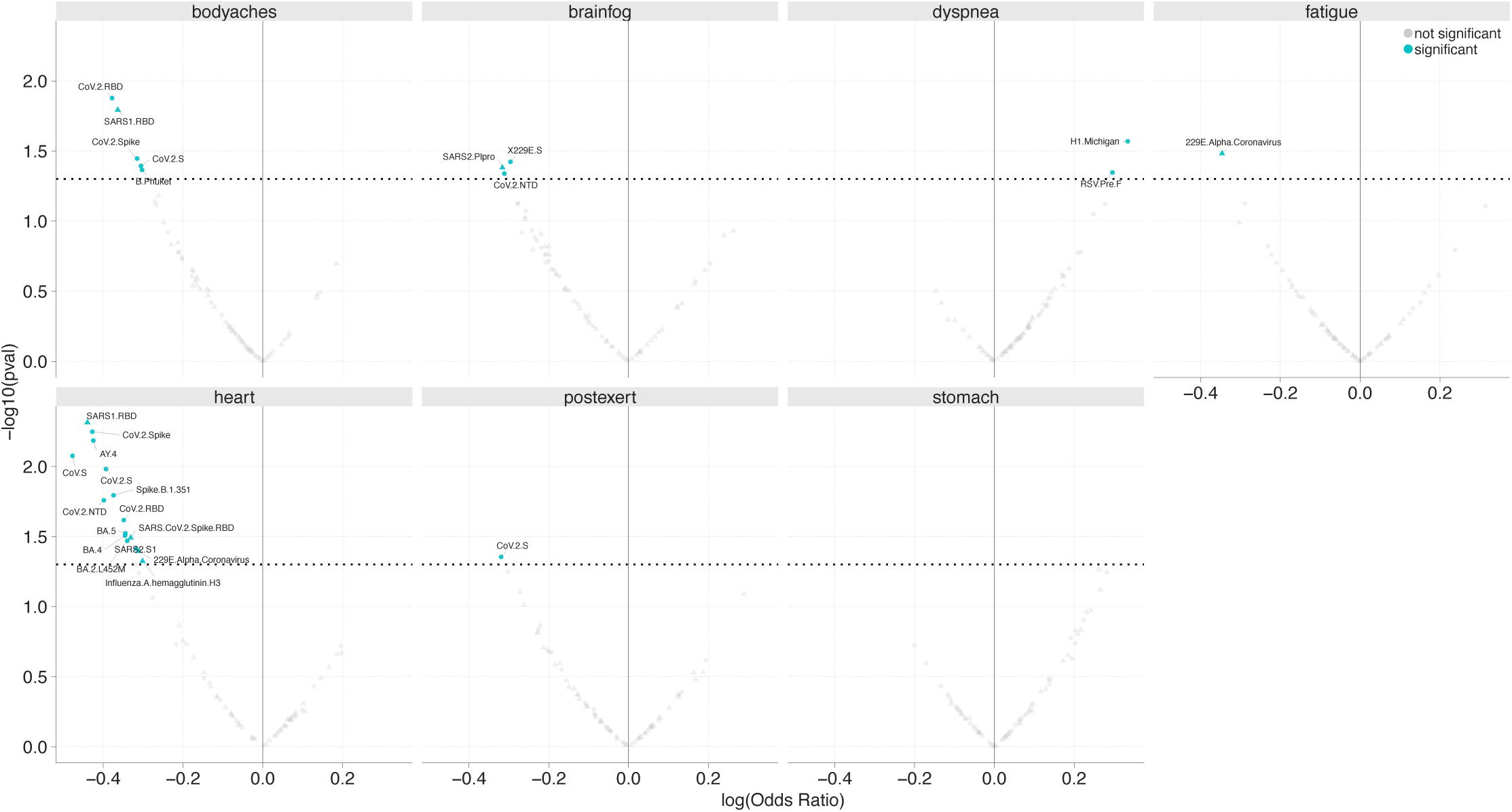
Association between antibody level and PRO severity in STOP-PASC at baseline as estimated by proportional odds logistic regression. The blue color indicates significance with *P*<0.05 (not corrected for multiple testing). Circles are measurements from MSD and triangles are from microbead assays.

**Figure S6.**
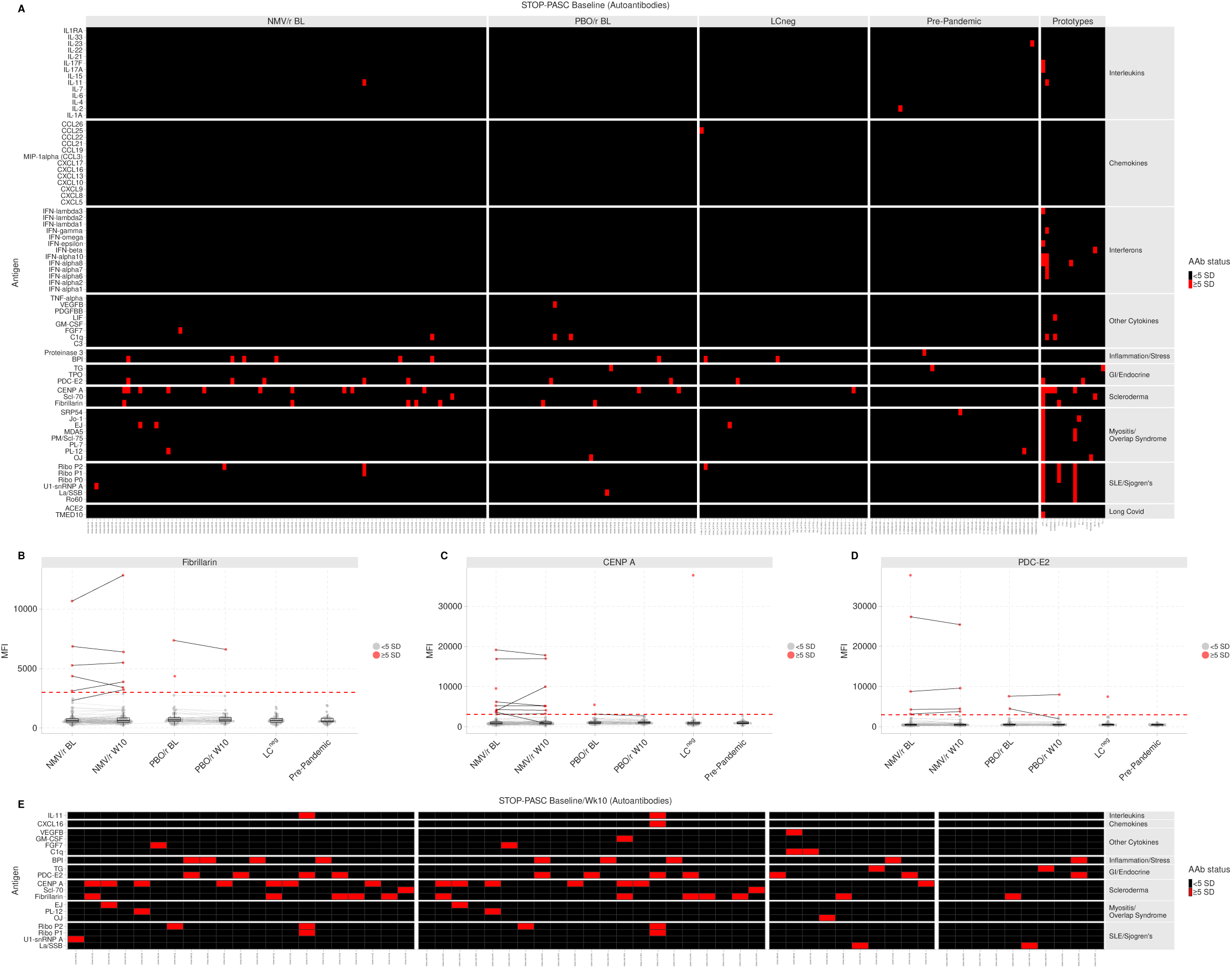
IgG autoantibody profiling of Long COVID patients. (A) Heatmap of autoantibody status from 76-plex bead-based protein array of cytokines, chemokines, growth factors, and receptors. Plasma of STOP-PASC Long COVID participants at baseline NMV/r (n=100), PBO/r (n=52), LCneg (n=42), healthy uninfected prepandemic controls (n=42), and prototype autoimmune disorders (n=16). Autoantigens are grouped based on disease (scleroderma, myositis, and overlap syndromes such as mixed connective tissue disease (MCTD), SLE/Sjögren’s, gastrointestinal and endocrine disorders), DNA-associated antigens, and antigens associated with tissue inflammation or stress responses. Colors indicate autoantibodies whose MFI measurements are >=5 SD (red) or <5 SD (black) above the average MFI for pre-pandemic controls. MFIs <3000 were excluded. (BD) Select autoantibody MFIs with a line connecting subjects at baseline and week10 in the NMV/r and PBO/r groups. (E) Autoantibody stability overtime at baseline (left box) and week10 (right box) for the set of Long COVID participants which had reactivities at baseline.

**Figure S7.**
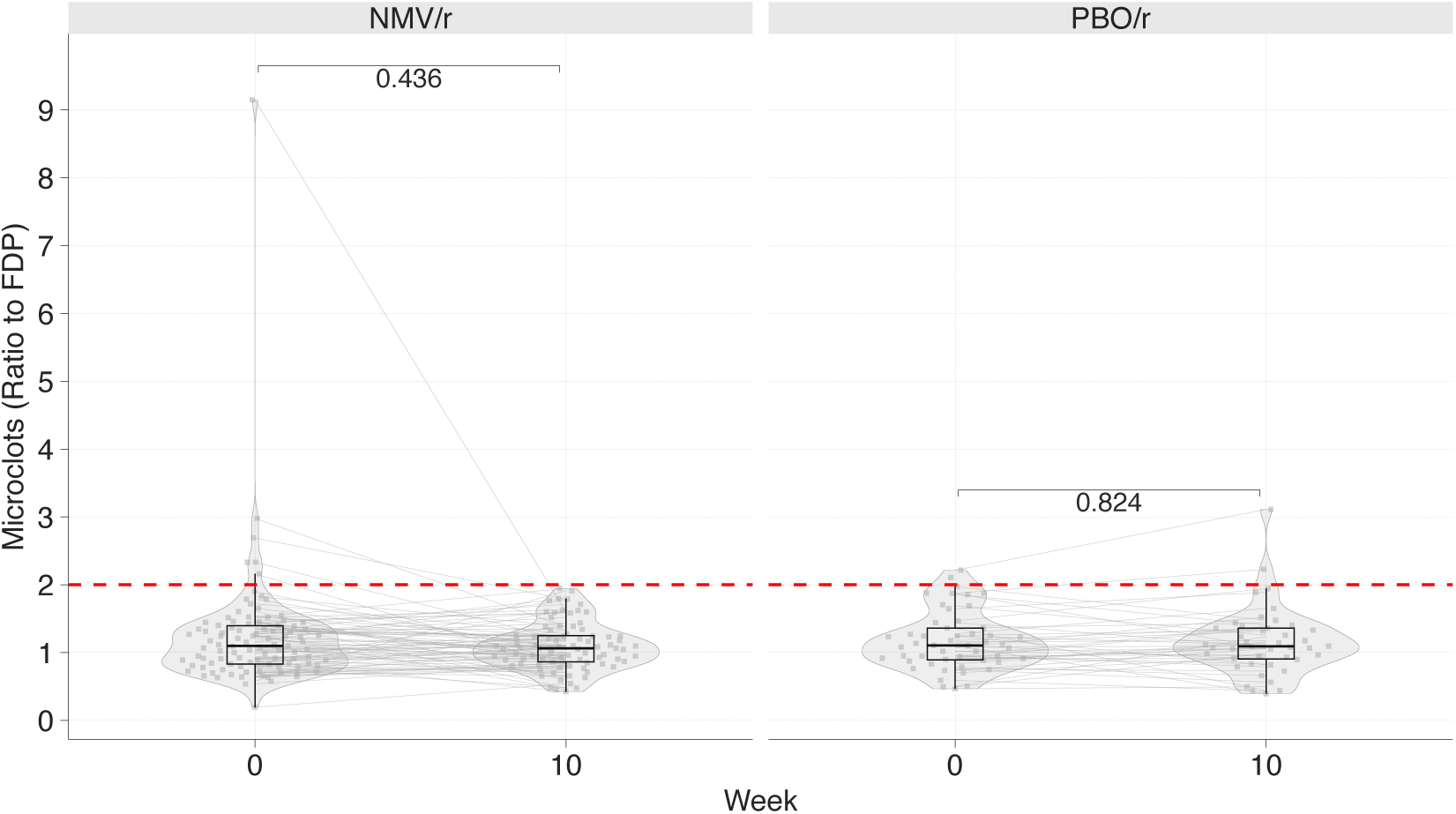
Microclotting assay at baseline and week10 in STOP-PASC LC participants.

**Figure S8.**
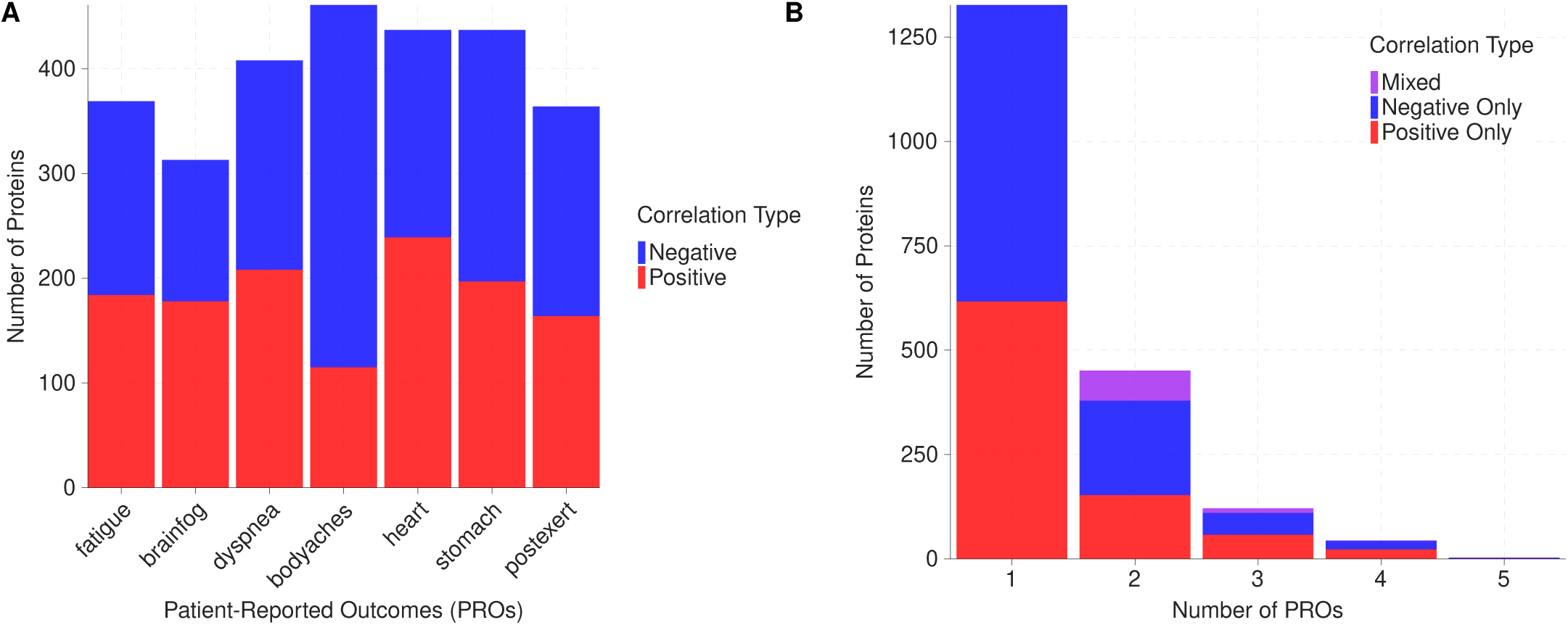
Protein correlations with patient-reported outcomes (PROs).

**Figure S9.**
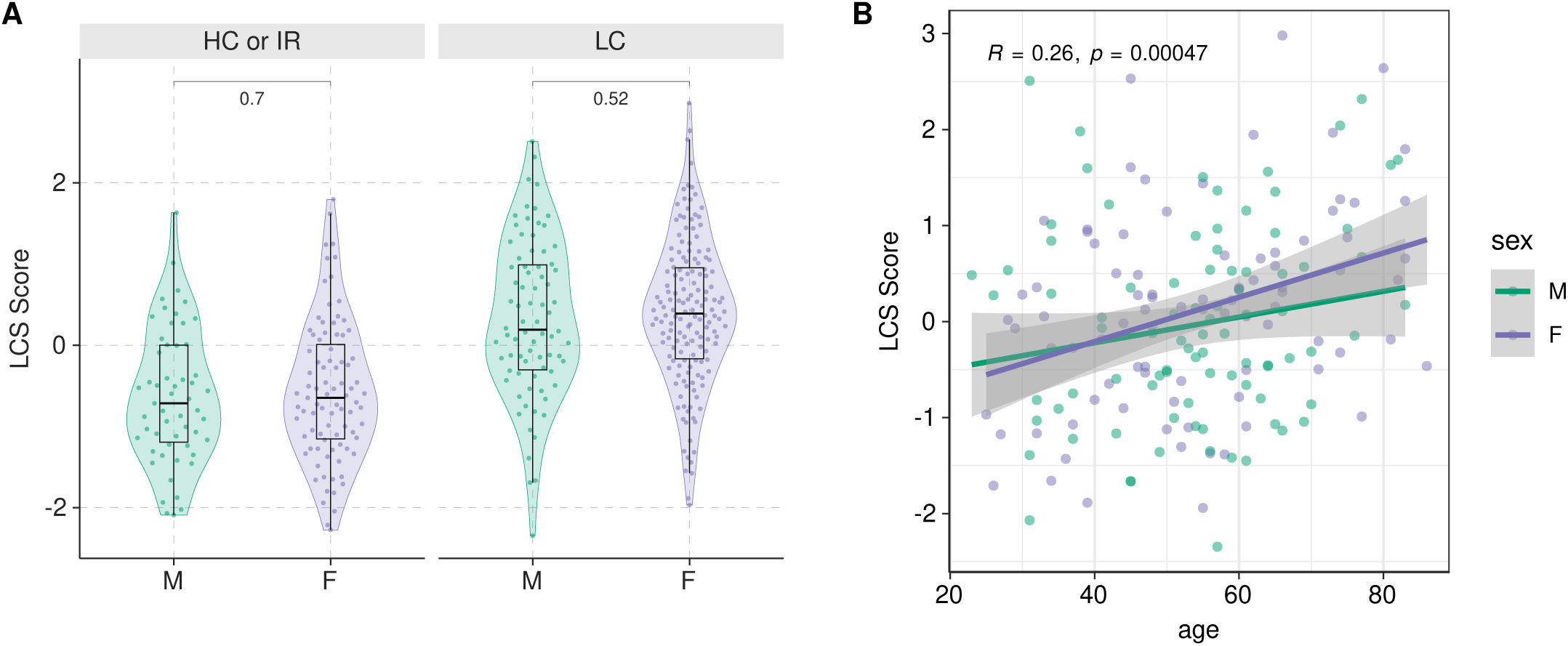
(A) Distribution of the LCS score by sex in Long COVID patients from public cohorts used in meta-analysis. The Wilcoxon rank-sum test was used for p-value calculation. (B) Spearman’s correlation between age and the LCS score.

**Figure S10.**
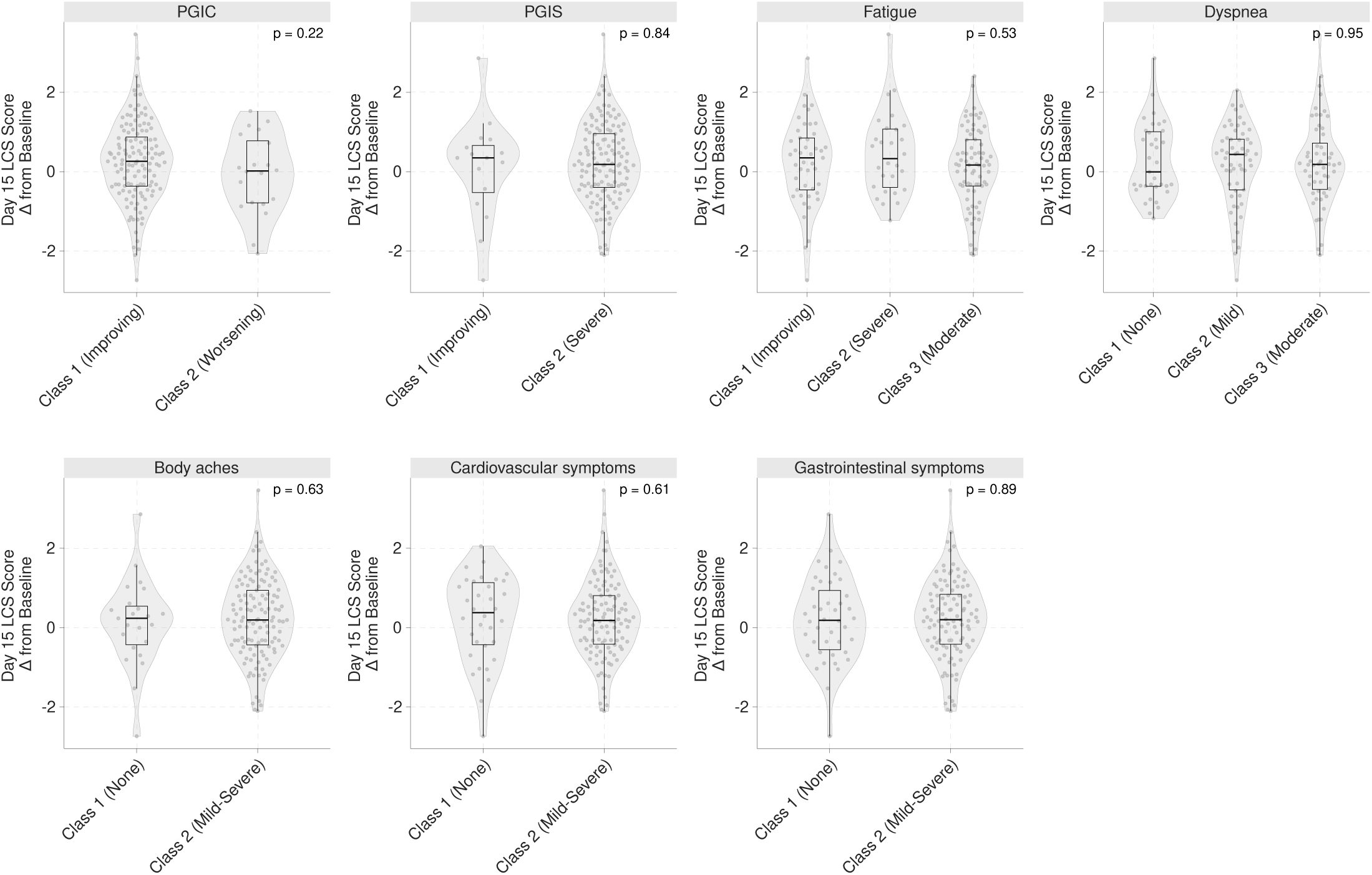
Day 15 change from baseline of the LCS Score in STOP-PASC participants comparing patient-reported outcome trajectory classes.

**Table S1.**
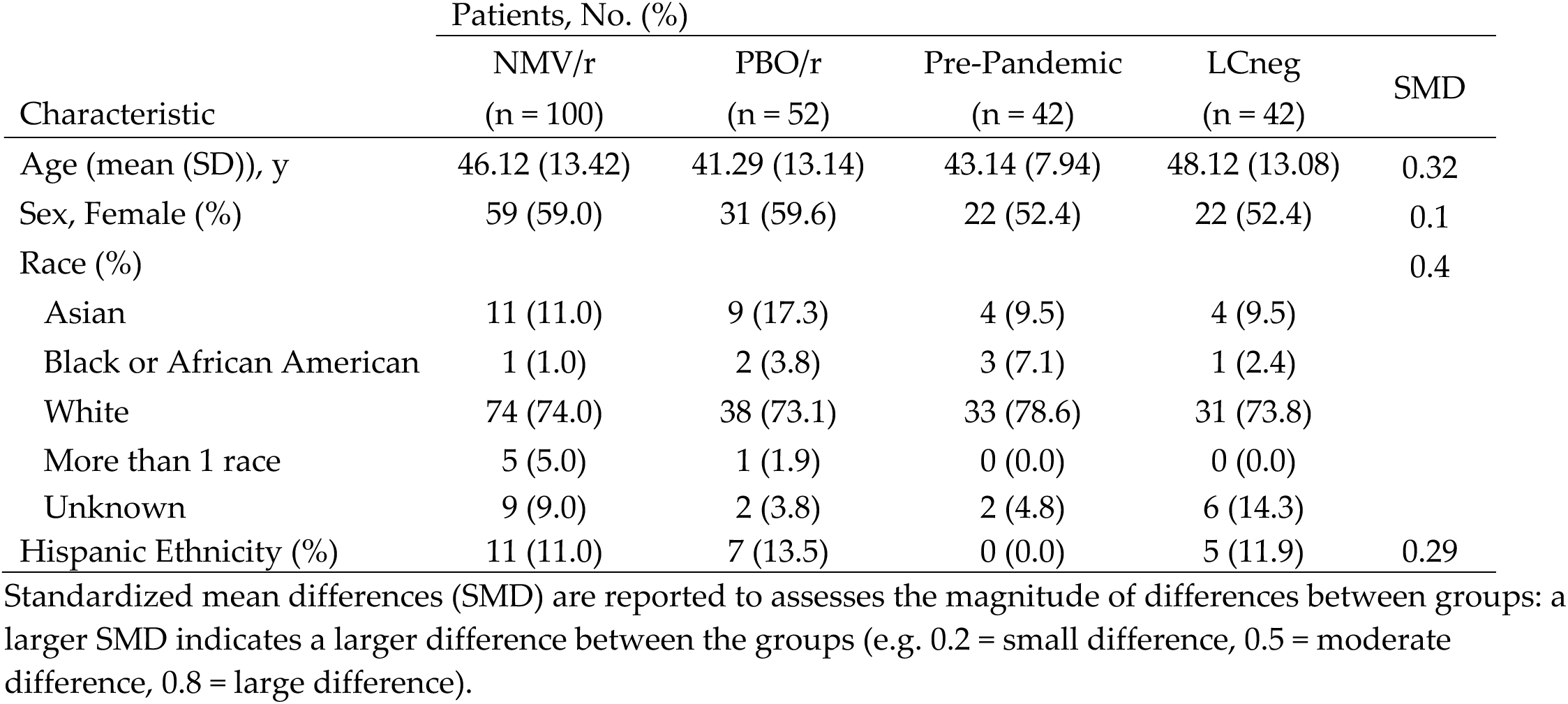
Characteristics of study populations.

| Characteristic | Patients, No. (%) |  |  |  | SMD |
| --- | --- | --- | --- | --- | --- |
|  | NMV/r<br>(n = 100) | PBO/r<br>(n = 52) | Pre-Pandemic<br>(n = 42) | LCneg<br>(n = 42) |  |
| Age (mean (SD)), y | 46.12 (13.42) | 41.29 (13.14) | 43.14 (7.94) | 48.12 (13.08) | 0.32 |
| Sex, Female (%) | 59 (59.0) | 31 (59.6) | 22 (52.4) | 22 (52.4) | 0.1 |
| Race (%) |  |  |  |  | 0.4 |
| Asian | 11 (11.0) | 9 (17.3) | 4 (9.5) | 4 (9.5) |  |
| Black or African American | 1 (1.0) | 2 (3.8) | 3 (7.1) | 1 (2.4) |  |
| White | 74 (74.0) | 38 (73.1) | 33 (78.6) | 31 (73.8) |  |
| More than 1 race | 5 (5.0) | 1 (1.9) | 0 (0.0) | 0 (0.0) |  |
| Unknown | 9 (9.0) | 2 (3.8) | 2 (4.8) | 6 (14.3) |  |
| Hispanic Ethnicity (%) | 11 (11.0) | 7 (13.5) | 0 (0.0) | 5 (11.9) | 0.29 |

Standardized mean differences (SMD) are reported to assess the magnitude of differences between groups: a larger SMD indicates a larger difference between the groups (e.g. 0.2 = small difference, 0.5 = moderate difference, 0.8 = large difference).

**Table S2.** Viral array content.

| Antigen | Vendor | Catalog # |
| --- | --- | --- |
| OC43 Beta Coronavirus | Peter Kim Lab | n/a |
| 229E Alpha Coronavirus | Peter Kim Lab | n/a |
| HKU1 Beta Coronavirus | Peter Kim Lab | n/a |
| SARS-CoV-2 Spike RBD | Sino | 40592-V08B |
| SARS ORF7A | ProSci | 10-435 |
| SARS ORF 8 | ProSci | 10-002 |
| SARS NSP8 | ProSci | 10-415 |
| SARS NSP9 | ProSci | 10-417 |
| SARS NSP12 | R&D | 10686-CV |
| SARS NSP13 | ProSci | 10-427 |
| SARS NSP15 | ProSci | 20-207 |
| SARS NSP16 | MyBioSource | MBS156017 |
| SARS NSP7 | Origene | TP750200 |
| SARS 3Clpro | Sino | 40594-V56E |
| SARS2 Plpro | Sino | 40593-V07E |
| SARS-CoV-2 Helicase | Sino | 40596-V07E |
| MERS RBD | Sino | 40071-V08B1 |
| TMED10 | n/a | n/a |
| EBV BFFL1 | n/a | n/a |
| EBV VCA | n/a | n/a |
| SARS1 RBD | Sino | 40150-V08B2 |
| Influenza A hemagglutinin (H1) | Peter Kim Lab | n/a |
| SARS-CoV-2 Methyltransferase | Sino | 40598-V07E |
| SARS-CoV-2 RDRP | Sino | 40595-V08B |
| SARS2 NSP10 | ProSci | 10-408 |
| SARS2 Spike S2 ECD | ProSci | 10-115 |
| Influenza A hemagglutinin (H3) | Peter Kim Lab | n/a |
| RSV-F Protein | Sino | 11049-V08B |
| EBV p18 | Prospec | ebv-277 |
| EBV EA-D | MyBioSource | MBS319448 |
| EBV EBNA-1 | Abcam | ab138345 |
| EBV EA | Prospec | EBV-272 |
| SARS2 ORF6 | ProSci | 20-193 |
| SARS2 S1 | ProSci | 97-087 |
| SARS2 Nucleocapsid | ProSci | 10-434 |
| SARS2 Envelope | ProSci | 97-082 |
| SARS2 Membrane | ProSci | 10-429 |
| SARS NSP3 | ProSci | 10-406 |
| HBsAg | MyBioSource | MBS142509 |
| Varicella Zoster Glycoprotein E | MyBioSource | MBS553197 |
| EV71 VP1 Capsid Protein | BioMART | Custom Request |
| Mumps Nucleoprotein | Prospec | MMP-001 |
| Rubella E1 | Prospec | RUB-291 |
| Measles Nucleoprotein | MyBioSource | MBS319759 |
| AcmNPV gp64 | Sino | 40496-V08B |
| Rhinovirus VP1 | MyBioSource | MBS1220686 |
| Parainfluenza Hemagglutinin-neuraminidase | Sino | 40629-V07B |
| HSV glycoprotein G | MyBioSource | MBS145445 |
| CMV gB | Prospec | CMV-211 |
| HMPV Glycoprotein G | Sino | 40791-V08H |
| Bare Bead | n/a | n/a |
| Anti-Human IgG Fc fragment Specific | Jackson | 109-005-008 |
| Anti-Human IgG F(ab') fragment specific | Jackson | 109-005-006 |
| Anti-Human IgG (H+L) | Jackson | 109-005-003 |
| Human IgG from serum | Jackson | I4506 |

**Table S3.**
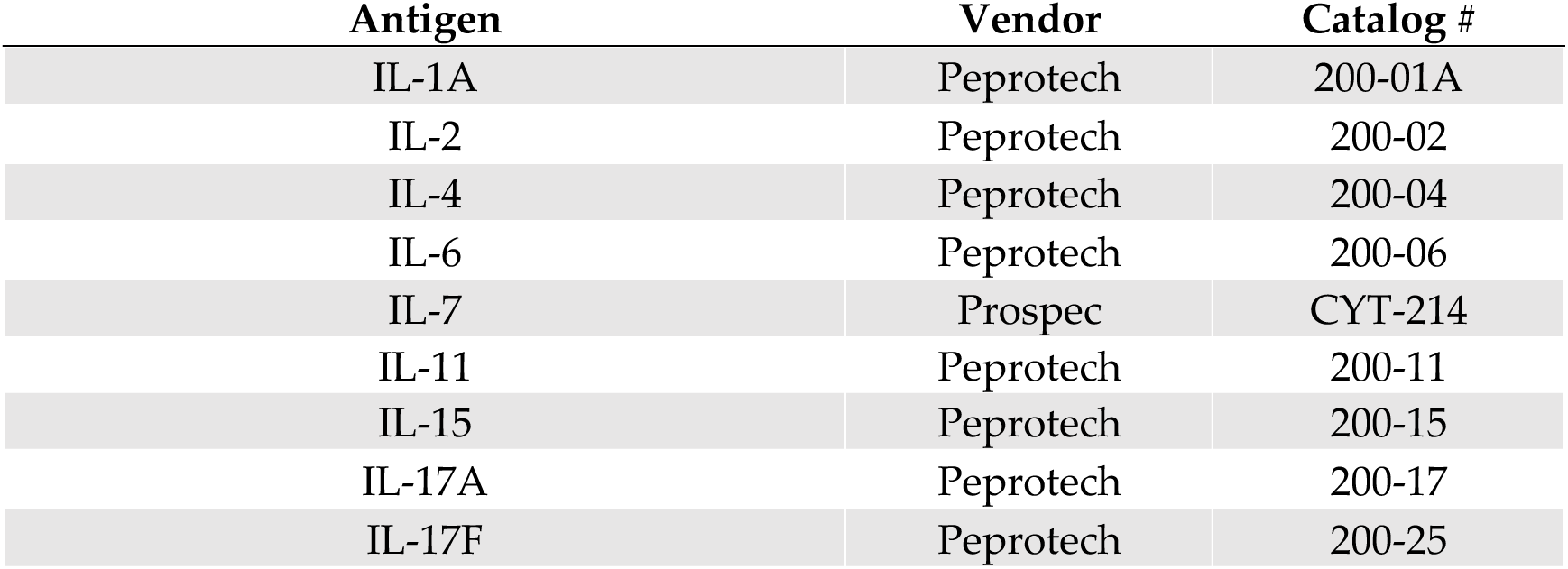
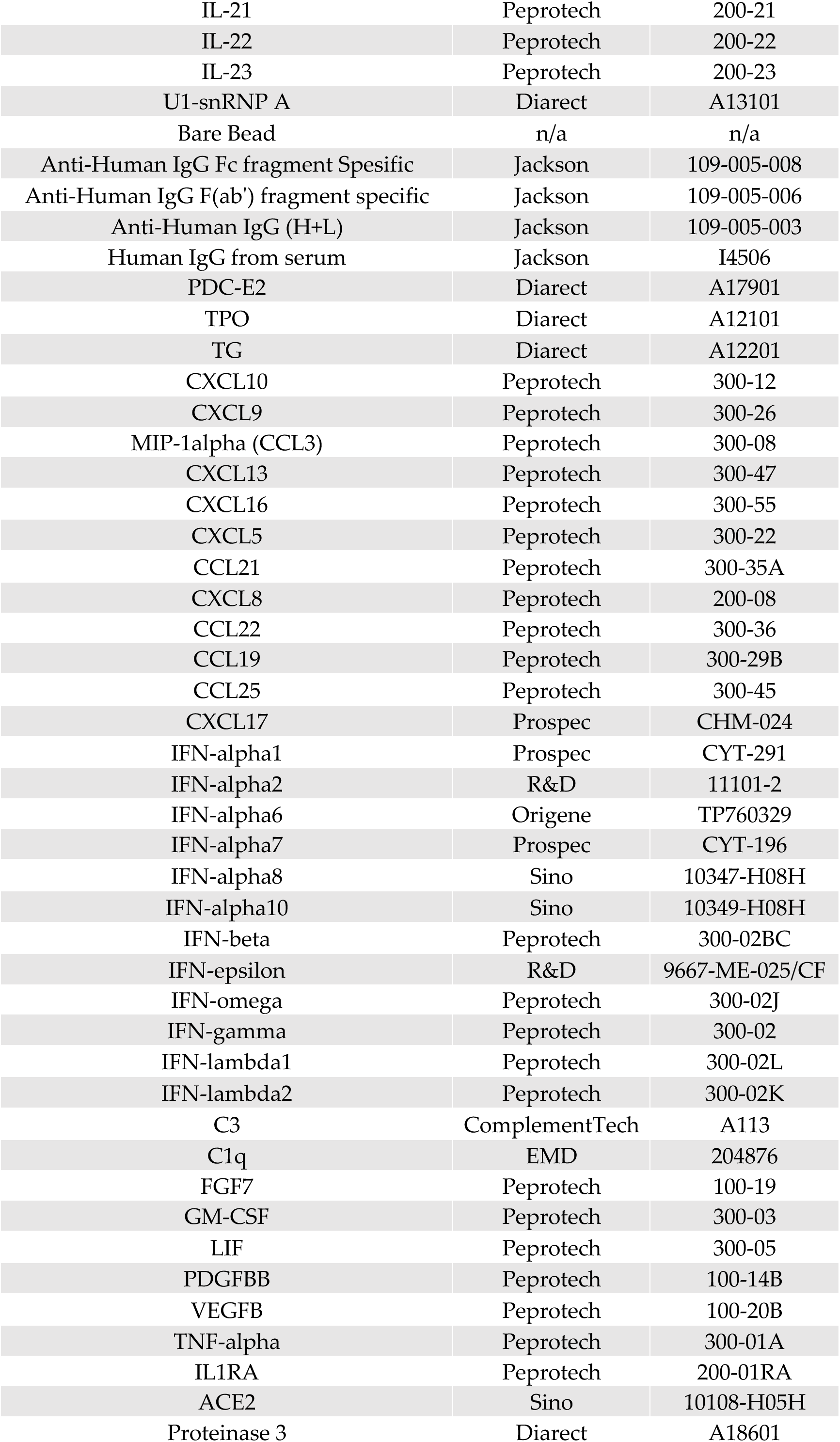
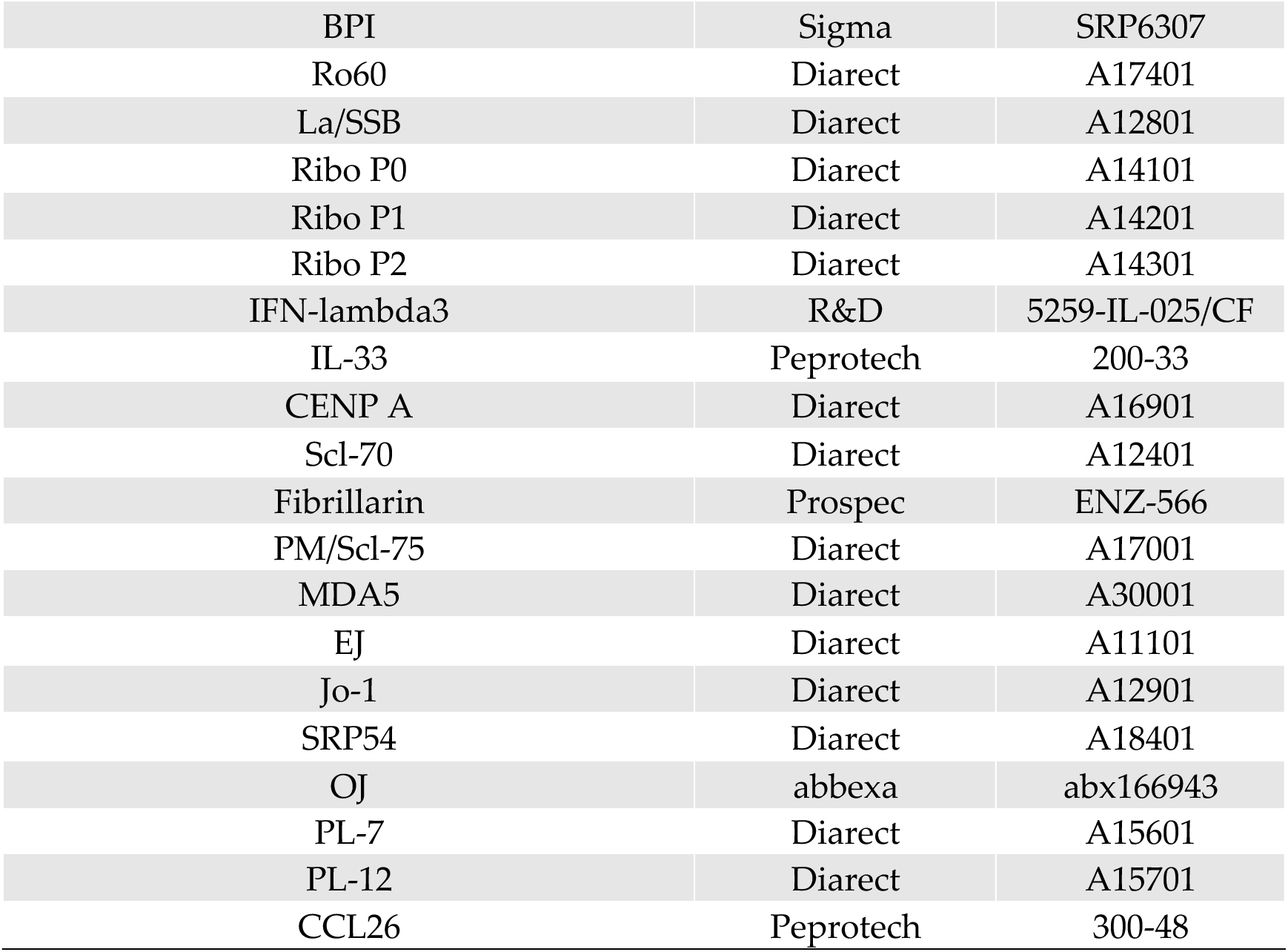
Luminex Autoantigen content.

**Table S4.**
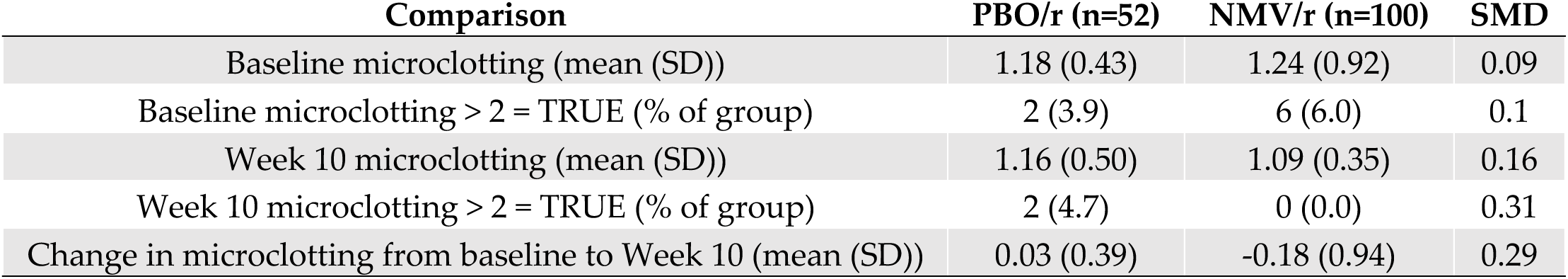
Microclotting results in STOP-PASC participants.

**Table S5.**
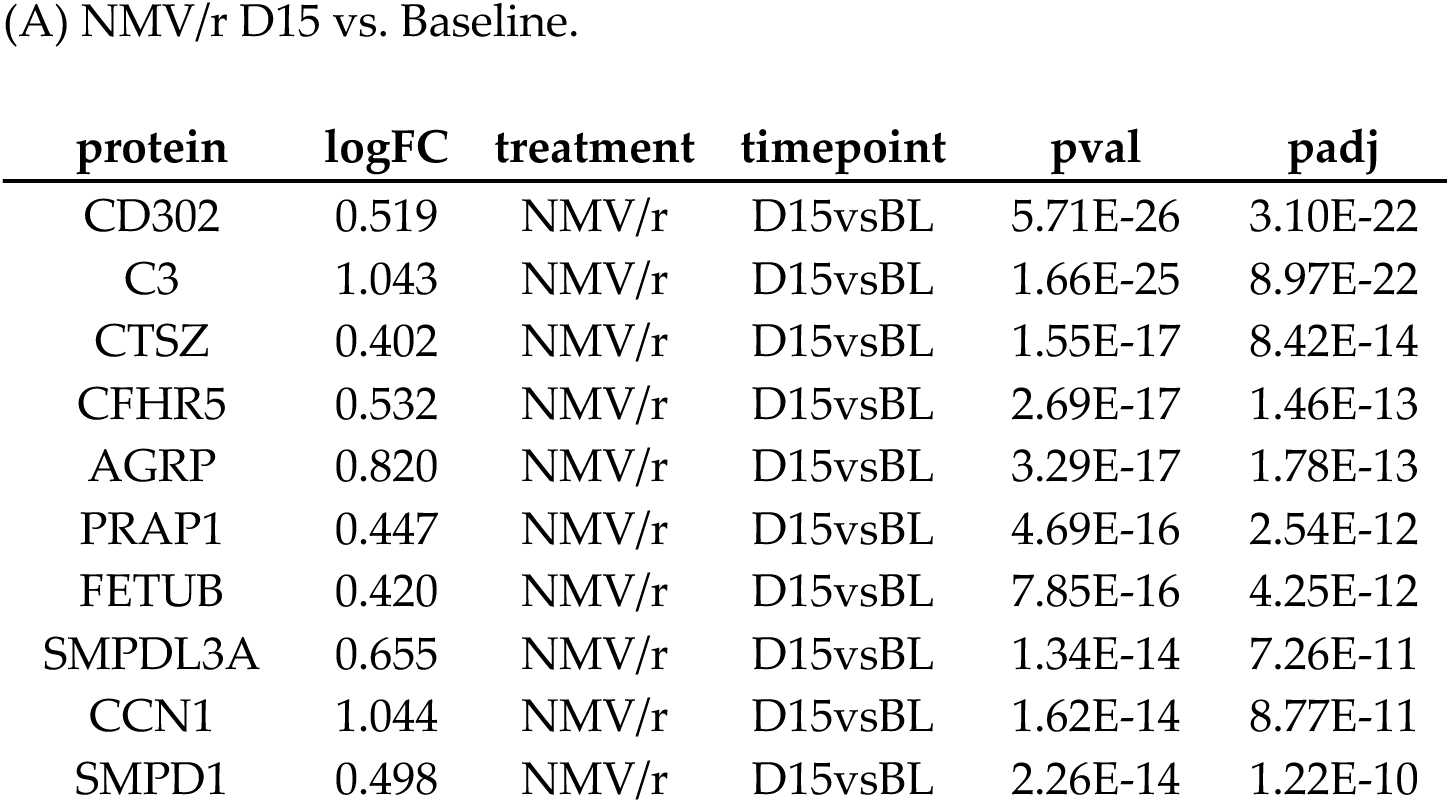
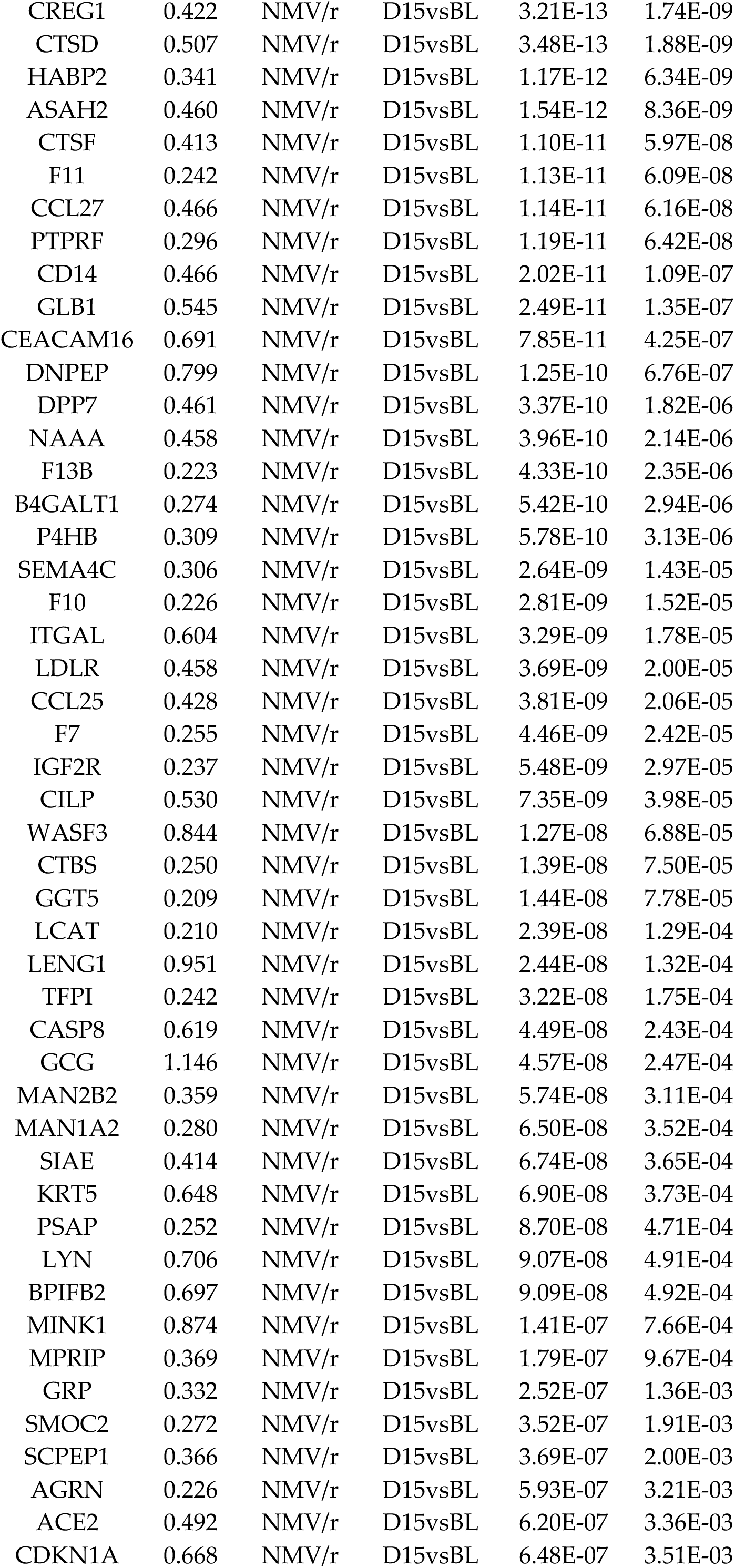
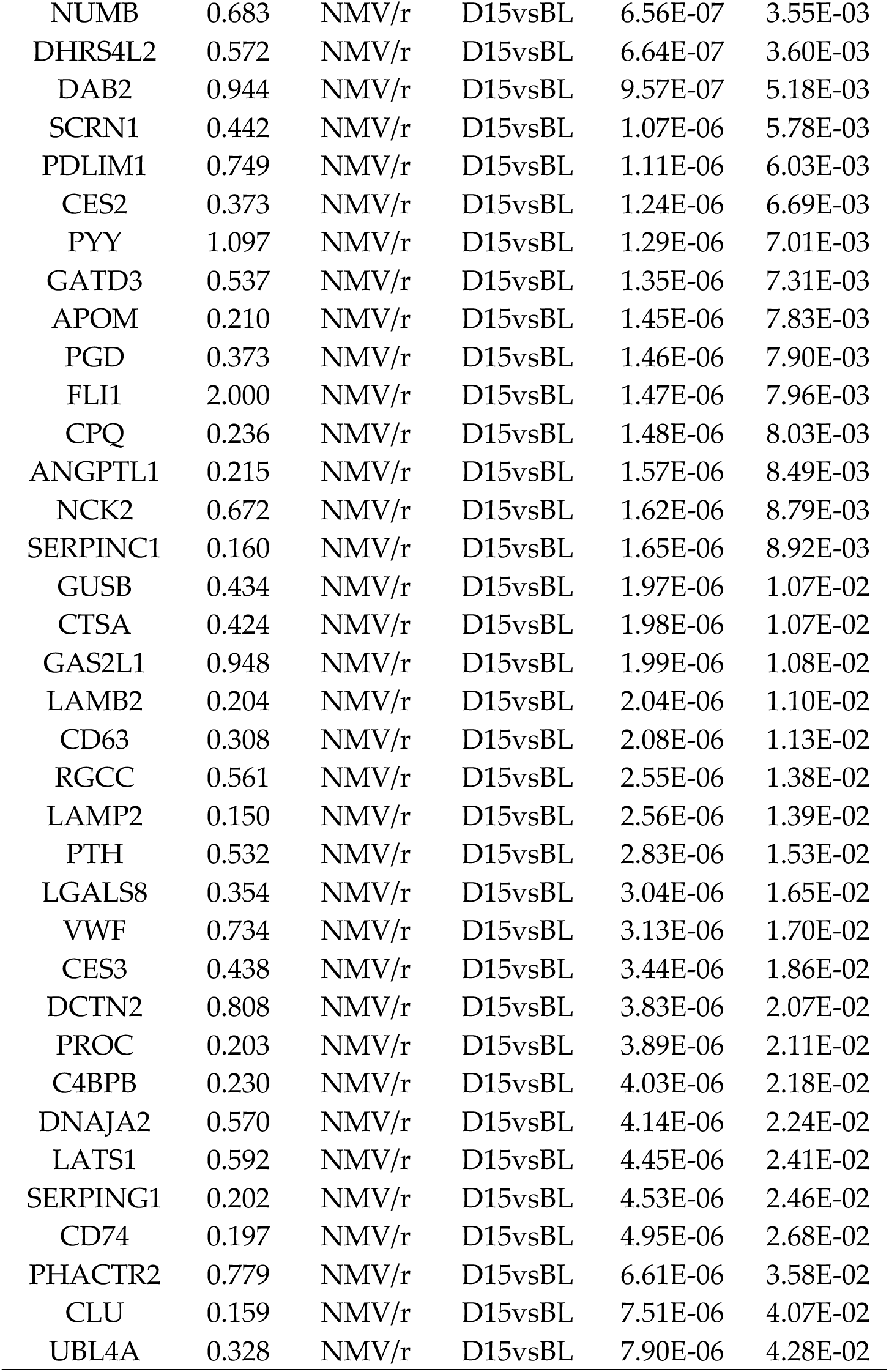
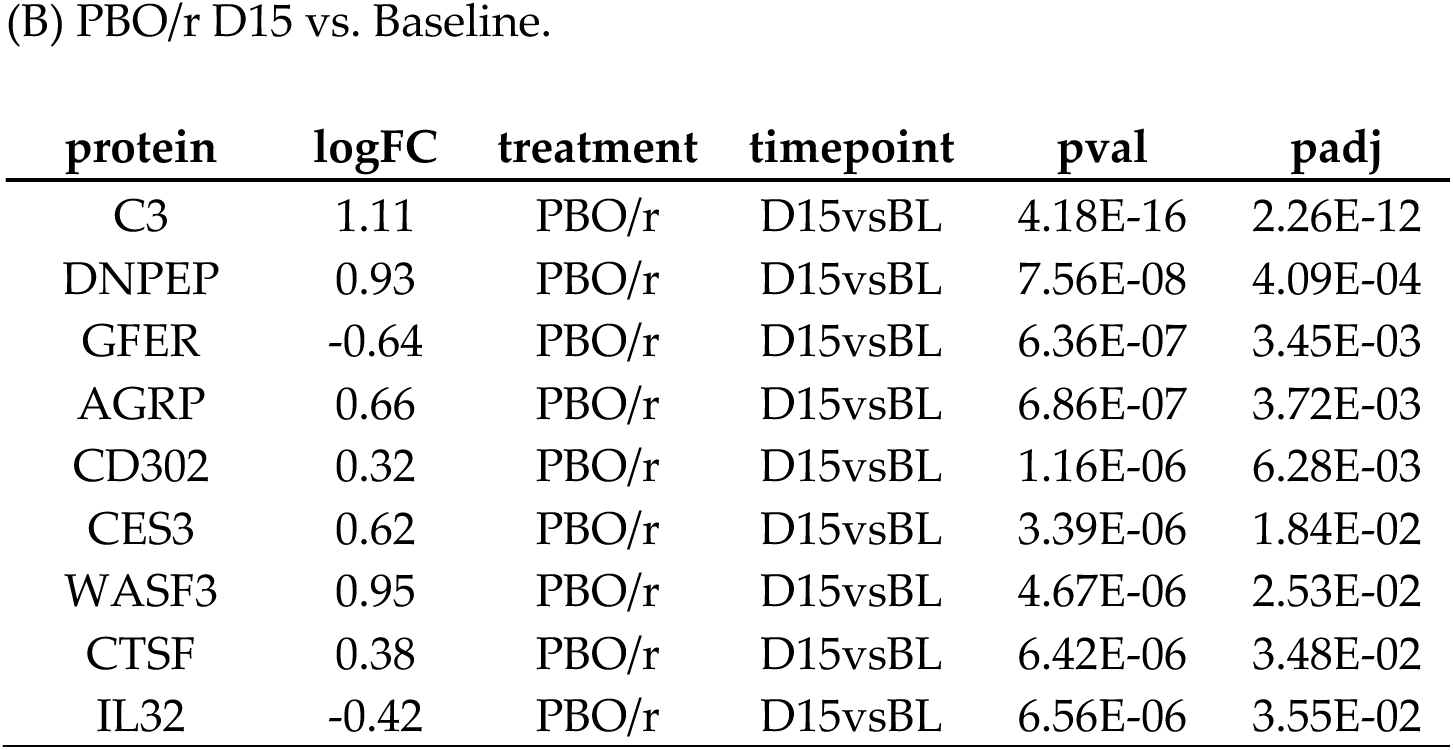
Differential proteins at D15 vs. Baseline in STOP-PASC participants.

**Table S6.** Publicly available Long COVID Olink cohort details.

| study | split | source | proteins<br>n | Long<br>COVID | Healthy Control<br>(HC) or Infected<br>Recovered (IR) | Time of Sample |
| --- | --- | --- | --- | --- | --- | --- |
| talla2023 | discovery | serum | 1463 | 26 | 22 (HC) 5 (IR) | symptoms lasting ≥60 days,<br>(max 379 DPSO) |
| mahdi2023 | discovery | plasma | 700 | 42 | 21 (HC) | ≈18 month time point post-<br>covid |
| vijayakumar2022 | discovery | plasma | 435 | 18 | 9 (HC) | symptoms lasting ≥ 3<br>months |
| su2022 | discovery | plasma | 442 | 43 | 30 (IR) | using ≥60 DPSO for LC |
| yin2024 | discovery | plasma | 384 | 25 | 15 (IR) | 8 month time point post-<br>covid |
| woodruff2023 | discovery | serum | 2925 | 92 | 26 (IR) | mean 140 DPSO, (22–446<br>DPSO) |
| fraser2023 | validation | plasma | 3072 | 22 | 22 (HC) | mean 104 DPSO, (48-168<br>DPSO) |
| rodriguez2024 | validation | plasma | 96 | 99 | 50 (IR) | ≥ 3 months |
| hamlin2024 | validation | plasma | 180 | 12 | 11 (IR) | using 12 month time point |

\*DPSO: Days Post Symptom Onset

| Discovery Total |  |
| --- | --- |
| Long<br>COVID | HC, IR |
| 246 | 52, 76 |
| Validation Total |  |
| Long<br>COVID | HC, IR |
| 133 | 22, 61 |

## Notes

### Clinical Trial

NCT05576662

### Author Declarations

Stanford University Institutional Review Board gave ethical approval for this work (clinicaltrials.gov NCT05576662).

